# Daily symptom monitoring is sustainable over months: retention, not compliance, is the primary barrier to long-duration digital tracking

**DOI:** 10.64898/2026.06.08.26355180

**Authors:** Chloe Z. Gunsilius, Pangzhongyuan Pei, Alexios Carayannopoulos, Frederike H. Petzschner

## Abstract

Ecological momentary assessment (EMA) enables real-time, longitudinal measurement of symptoms and behavior via smartphones, yet nearly all feasibility evidence comes from protocols lasting one to two weeks, far shorter than the timescales over which chronic diseases fluctuate and clinical decisions unfold. Whether daily compliance can be sustained over months, or whether it decays as short-protocol trends predict, is unknown. Here, 214 participants (173 with pain, 41 healthy controls) completed a 4-month (122-day) EMA protocol via the Soma smartphone app, generating 26,907 check-ins. Half the sample completed the full protocol without a two-week lapse. Aggregate compliance appeared moderate (50%), but this conflated two distinct phenomena: when recomputed over each participant’s active period, compliance rose to 71%, with 91% achieving moderate-to-high adherence, and remained stable across all 17 study weeks. Pain status predicted earlier disengagement but not lower compliance among those who remained; after adjustment for differential retention, group differences disappeared. To our knowledge, this is the longest continuous daily EMA evaluation in a clinical population. It suggests the primary barrier to long-duration EMA is not declining motivation among active participants but concentrated early disengagement, with direct implications for the design of digital health protocols, decentralized trials, and remote symptom monitoring.

## Introduction

Much of what we know about how people feel, think, and cope in daily life comes from asking them to remember. Retrospective questionnaires, administered at clinic visits or research appointments weeks or months apart, compress continuous, fluctuating experience into a handful of static snapshots. This introduces recall bias, strips away temporal dynamics, and divorces measurement from the context in which experience actually occurs (Shiffman, Stone, and Hufford 2008). The distortion is not random: retrospective reports diverge from real-time experience in structured, predictable ways, shaped by cognitive biases including peak-end effects and duration neglect (Redelmeier and Kahneman 1996; Schwarz 1999; Shiffman, Stone, and Hufford 2008; Stone et al. 2004).

In chronic disease, where objective biomarkers are scarce for many of the symptoms that matter most, this problem is compounded. In this case, patient-reported outcomes (PROs) are among the most common endpoints for gauging treatment efficacy, guiding clinical decisions, and determining trial results (Calvert et al. 2013; U.S. Department of Health and Human Services et al. 2009). Yet PROs remain overwhelmingly retrospective in practice (Basch 2010; Calvert et al. 2018). For symptoms that fluctuate continuously, like pain, fatigue, and mood, a questionnaire administered every few weeks may not reliably reflect what patients actually experienced. The within-person trajectories that matter most for clinical care, including how symptoms evolve day to day, how they respond to treatment, or whether a flare is building, are precisely what retrospective snapshots cannot capture.

Ecological momentary assessment (EMA) was developed to address precisely this gap. Rather than relying on participants to reconstruct their experience from memory, EMA samples what is happening during ordinary daily life, close to the moments that matter (Larson and Csikszentmihalyi 2014; Shiffman et al. 1997; Shiffman, Stone, and Hufford 2008; Stone et al. 2004). With the widespread adoption of smartphones, the method has become scalable in ways its originators could not have anticipated, enabling dense longitudinal data collection from large samples without constant researcher oversight (Wrzus and Neubauer 2023). The clinical value of this approach is now well documented across domains: within-person temporal dynamics captured by EMA have revealed diagnostic and prognostic signals that cross-sectional snapshots cannot detect (Ebner-Priemer and Trull 2009; Mun et al. 2019; M. Wichers 2014; Marieke Wichers et al. 2012), from relapse triggers (Shiffman et al. 1997) to moment-to-moment precursors of depressive episodes (Moore et al. 2016).

The same properties that give EMA its scientific value now place it at the center of digital health. Electronic PRO platforms are increasingly integrated into clinical trials (Basch et al. 2016; Calvert et al. 2018), systematic symptom self-monitoring has been associated with improved quality of life and longer survival in cancer care (Basch et al. 2017; Denis et al. 2019), and continuous patient-generated data are increasingly positioned to enable personalized treatment adjustment, early detection of deterioration, and more sensitive trial endpoints (Coravos, Khozin, and Mandl 2019; Dorsey et al. 2017). However, the infrastructure of remote monitoring, decentralized trials, and digital therapeutics rest on one assumption: that patients will continue to report their experience, reliably, over weeks and months. Whether they will is an assumption under-tested in the field.

Almost everything we know about EMA feasibility comes from studies that last about a week. The largest cross-field meta-analysis found a mean duration of 7 days with an average compliance rate of 79% (Wrzus and Neubauer 2023); studies extending beyond two weeks are uncommon (May et al. 2018; Ono et al. 2019), and none of the 12 chronic pain EMA datasets pooled by Ono et al. (2019) exceeded 28 days. Yet the clinical questions that motivate long-duration monitoring, including how symptoms fluctuate over months, how interventions affect long-term outcomes, and how acute conditions become chronic, require observation windows measured in months rather than days (Cohen, Vase, and Hooten 2021). The field lacks the empirical feasibility evidence it needs to get there.

Whether long-duration EMA is achievable remains an open question, and the available evidence offers little reassurance. Declining engagement is among the most consistent observations in mobile health research, with pooled dropout rates approaching 43% in app-based chronic disease interventions (Amagai et al. 2022; Eysenbach 2005; Meyerowitz-Katz et al. 2020). Dropout is rarely random: the participants who leave earliest are often those whose data the study most needs, introducing survivorship bias that can distort conclusions (Eysenbach 2005). In clinical populations, symptom burden, fatigue, and fluctuating motivation may compound these pressures in ways that short studies of healthy volunteers cannot anticipate (Jamison et al. 2017). The short-protocol evidence we do have is not reassuring when projected forward: the largest individual-patient-data meta-analysis of EMA in chronic pain found that completion rates declined by roughly 2.3 percentage points per week over studies lasting 4 to 28 days (Ono et al. 2019). Extrapolated to a four-month protocol, that trajectory would drive compliance to near zero by the third month. Whether compliance actually follows this trajectory, or stabilizes once participants settle into a routine, cannot be determined from protocols that end before the answer would become visible. For that, EMA needs to scale in duration.

Chronic pain offers a particularly informative proving ground for testing long-duration EMA. Pain affects more than 20% of U.S. adults (Dahlhamer et al. 2018) and is among the most prevalent transdiagnostic symptoms across chronic disease, routinely tracked as a PRO endpoint in oncology, rheumatology, neurology, and cardiovascular medicine (Dworkin et al. 2005; Kluetz et al. 2016; Treede et al. 2019). Pain is inherently dynamic: within-person fluctuations are substantial and clinically meaningful, and the trajectory of pain over weeks and months sits at the center of treatment decisions (Cohen, Vase, and Hooten 2021; Mun et al. 2019). Testing whether daily EMA can be sustained over months in a mixed sample of individuals with pain and healthy controls therefore allows two questions to be addressed simultaneously: whether living with a chronic, fluctuating condition constitutes a barrier to sustained EMA participation, and whether the resulting feasibility benchmarks generalize beyond pain research.

Here, we report what we believe to be the longest continuous evaluation of intensive daily EMA in a clinical population: a 122-day prospective protocol in which 214 participants (173 with acute or chronic pain, 41 healthy controls) generated about 27,000 EMA observations using the Soma smartphone app (Gunsilius et al. 2024). Participants completed up to three EMA-style check-in types each day, including scheduled evening assessments, pseudo-randomly prompted daytime momentary assessments, and self-initiated voluntary check-ins.

We characterize engagement, prompted compliance, and retention across the full 122-day window, providing the empirical feasibility benchmarks the field currently lacks for multi-month EMA. Central to this is a distinction that existing studies rarely make explicit, which is to separate two dimensions of adherence that are often conflated: *retention*, whether participants remain in the protocol at all, and *compliance*, how reliably retained participants respond to prompts. These tell fundamentally different stories about feasibility in the long-duration setting. We show that compliance among retained participants does not decay over time, challenging the assumption that longer protocols inevitably produce thinner data. We compare clinical and non-clinical populations directly, finding that pain status predicts dropout but not the engagement of those who stay, and that once compliance is adjusted for differential retention, the group difference disappears. We also test whether dropout can be predicted from baseline characteristics and first-week behavior, identifying early signals that could inform targeted re-engagement. Nearly half of participants completed the protocol without a single two-week lapse, and among those who remained active, neither check-in intensity nor prompted compliance showed any measurable decline from the first week to the last. These results have direct implications for the design of long-duration digital health protocols, decentralized trials, and remote patient monitoring across clinical domains.

## Methods

### Study Design and Setting

This was a prospective, observational study in which eligibility screening, baseline questionnaire assessment, and EMA data acquisition were conducted entirely remotely. A subset of participants also completed additional in-laboratory measurements at Brown University; their EMA and questionnaire data are included in the present analyses, but the laboratory measurements are not reported here. Participants were recruited across the United States between June 2023 and March 2026 through three channels: community advertisements — physical posters displayed in coffee shops, libraries, and other public locations across Rhode Island; as well as online nationwide platforms, primarily Facebook, Google Ads, and email listservs; and clinical referrals from two local sites.

Eligible individuals were enrolled in a four-month (122-day) ecological momentary assessment protocol delivered through the Soma smartphone app (Gunsilius et al. 2024). Depending on the months of enrollment, the four-month period ranged from 120-122 days. To be conservative, we used 122 days for everyone. The study was purely observational; no intervention was administered as part of the study, though participants could indicate what treatments or interventions they utilized as part of their usual care within the daily Soma app check-ins. Following an online eligibility screening, participants provided informed consent and completed a baseline battery of self-report questionnaires (T0) assessing demographics, pain characteristics, pain catastrophizing, and psychological functioning. They were then invited by email to download the in-house developed pain tracking application (“SOMA Pain Manager”), available on both the Apple App Store and Google Play Store, and to register as a study participant to begin the EMA portion of the protocol. All online eligibility, consent, and questionnaires were delivered using the Qualtrics platform.

The study was approved by the Brown University Institutional Review Board (Protocol No. 2022003301). All participants provided written informed consent in accordance with the Declaration of Helsinki (2013). This study was pre-registered as an observational study on clinicaltrials.gov (NCT05754190) in March 2023.

### Participants and Recruitment Flow

To support recruitment of a sample reflective of real-world smartphone users with pain, eligibility criteria were intentionally kept broad. Rather than restricting enrollment by pain etiology, comorbidity profile, concurrent treatment, or prior digital health experience, we imposed only those exclusions necessary to ensure participants could feasibly complete the 122-day protocol. This real-world sampling approach was adopted to maximize the external validity of feasibility benchmarks derived from the cohort and to better approximate the heterogeneity of populations likely to be enrolled in future remote, app-based studies. Eligibility required age ≥18 years, ownership of a personal smartphone with stable internet access, and sufficient English fluency to complete study assessments. Exclusion criteria included logistical barriers to sustained participation, inability to commit to the four-month protocol, a current diagnosis of primary or metastatic cancer, and failure to meet the criteria for any of the three study cohorts. Participants were recruited into one of three groups: chronic pain (CP), acute/subacute pain (AP), or healthy control (HC) see the next section.

Between June 2023 and March 2026, 2,943 individuals completed an online eligibility screening. Of those screened, 429 provided informed consent and 341 completed the baseline questionnaire battery (T0). After removing suspicious and duplicate accounts (n = 12), we had a valid T0 sample of 329 participants. Out of those who completed T0, 256 participants decided to enroll in the Soma app to take part in the EMA tracking part of the study (73 did not enroll), whereby a subset of those still took part in a local in person part of the study (not reported here). One additional participant was excluded because the gap between T0 completion and app enrollment exceeded 40 days (n = 1), yielding a cleaned sample of 255 EMA participants.

Soma is an ongoing study with continuous, rolling enrollment; participants join individually and complete the 122-day protocol on their own timeline. The present paper reports on the subset who had completed the full observation window as of the analysis cutoff date (March 16, 2026). Thirty-seven participants who were still actively completing the protocol at that date were excluded to ensure that every included participant had an equal opportunity to contribute the full 122-day window (N = 218). A further four participants who enrolled but never submitted a single check-in were excluded from the analytic sample, as they provide no EMA engagement data (final analytic N = 214; Figure 1). 27 participants explicitly withdrew at some point during the 4-month app period by contacting study staff via phone or email to say they were withdrawing from the study. To allow a full picture of the feasibility data set, these participants who explicitly withdrew are included in the analytic sample and assessed in more depth.

**Figure 1.**
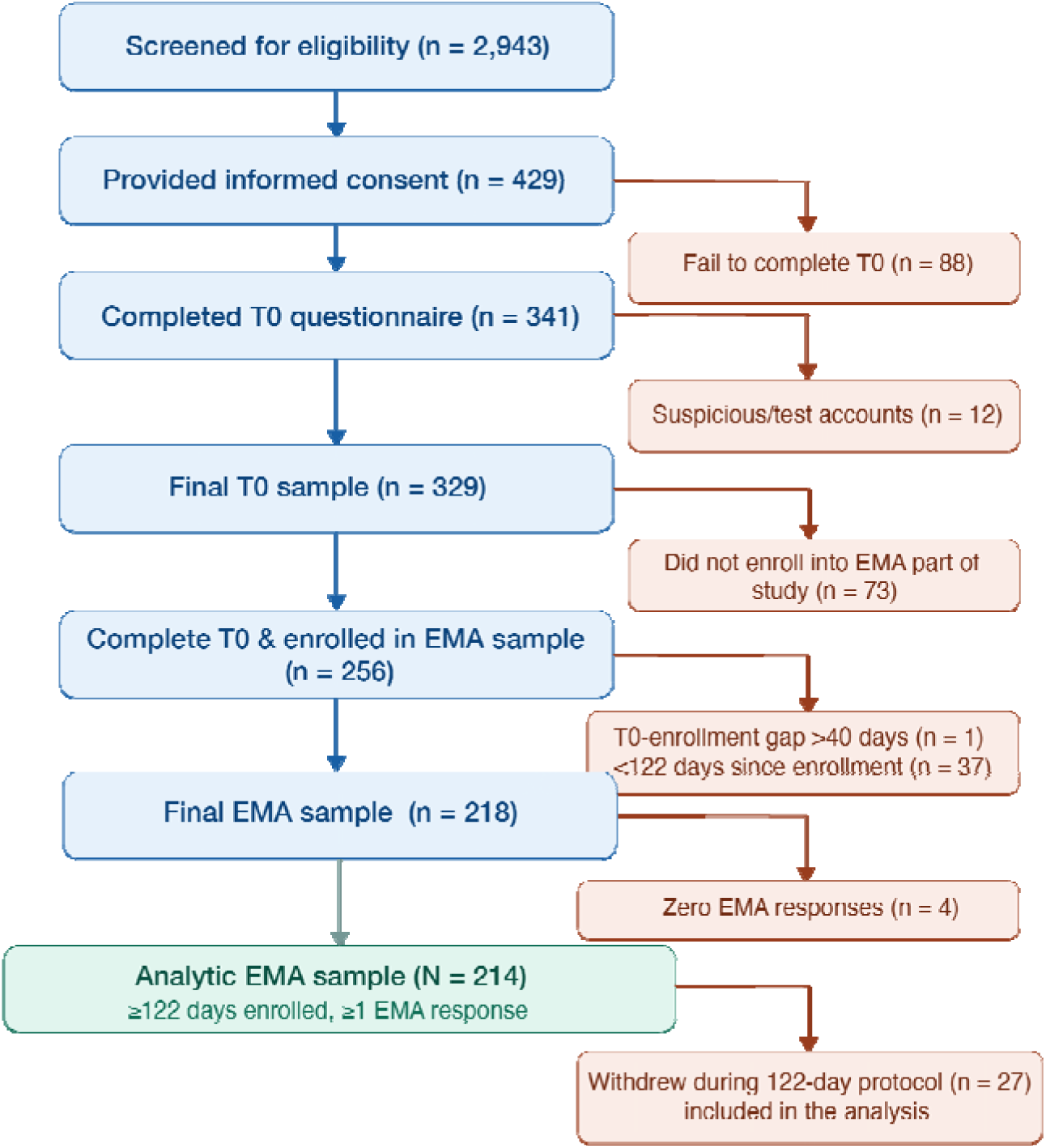
Participant flow diagram. Flow from eligibility screening (n = 2,943) to the Paper 1 analytic sample (N = 214). Exclusions at each step are shown to the right.

### Pain Group Classification

Participants were assigned to one of three pain groups, Healthy Control, Acute/Subacute Pain, or Chronic Pain, based on responses to the eligibility screening questionnaire. The classification algorithm used five items: current regular pain duration, current irregular pain duration, and three severity items rated on a 0–10 numerical rating scale (pain intensity, pain interference, and pain-related distress). A severity threshold of ≥3 on any of the three items was used as the criterion for clinically relevant pain. This threshold reflects the lower bound of the mild/moderate boundary established for the 0–10 NRS in non-cancer chronic pain populations (Boonstra et al. 2016; Hoffman et al. 2010) and follows the general framework of grading pain severity by its functional interference. It was deliberately set at the inclusive end of the published range, consistent with the study’s broad-eligibility design.

Group assignment proceeded as follows. Participants were classified as Chronic Pain if they reported either regular or irregular pain lasting more than six months and met the severity threshold (any rating ≥3). Participants were classified as Acute/Subacute Pain if their pain duration was six months or less on either item and they met the severity threshold. Participants were classified as Healthy Controls if they reported no or short-duration (<3 months) current regular or irregular pain and did not meet the severity threshold (all ratings <3). The final analytic sample comprised 214 participants: 41 Healthy Controls, 19 Acute/Subacute Pain, and 154 Chronic Pain. The relatively small Healthy Control group reflects the study’s primary recruitment focus on individuals living with pain, consistent with Soma’s design as a pain monitoring platform; healthy controls were recruited as a feasibility comparison group rather than as a matched sample, and the resulting imbalance is acknowledged in the Discussion. For all primary feasibility analyses, the Acute/Subacute and Chronic Pain groups are combined into a single Pain group (n = 173) and contrasted with Healthy Controls (n = 41) (**Table 1**). This two-level grouping reflects the primary feasibility question, whether individuals living with pain engage differently with a long-term EMA protocol than pain-free controls, rather than distinctions between pain subtypes.

**Table 1.**
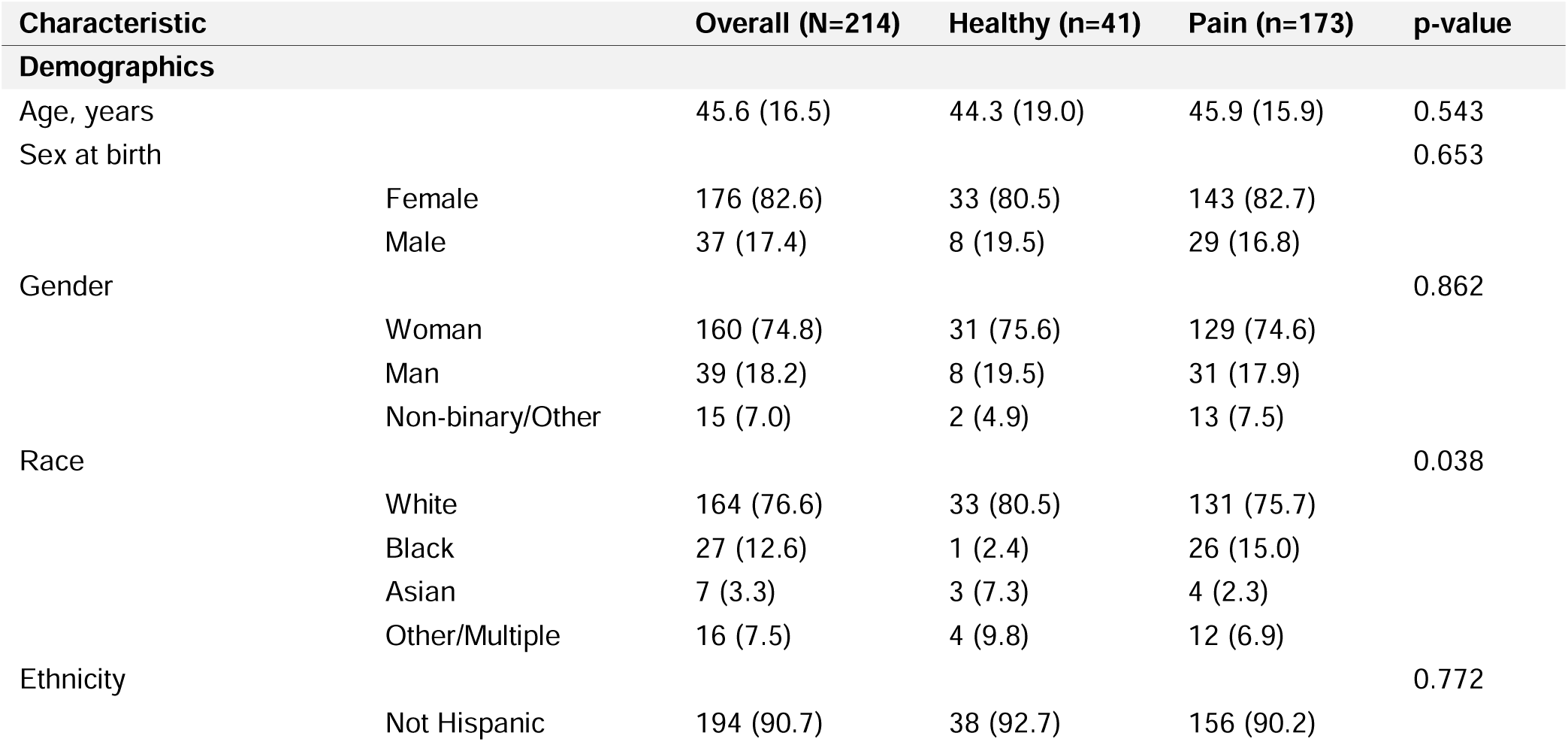
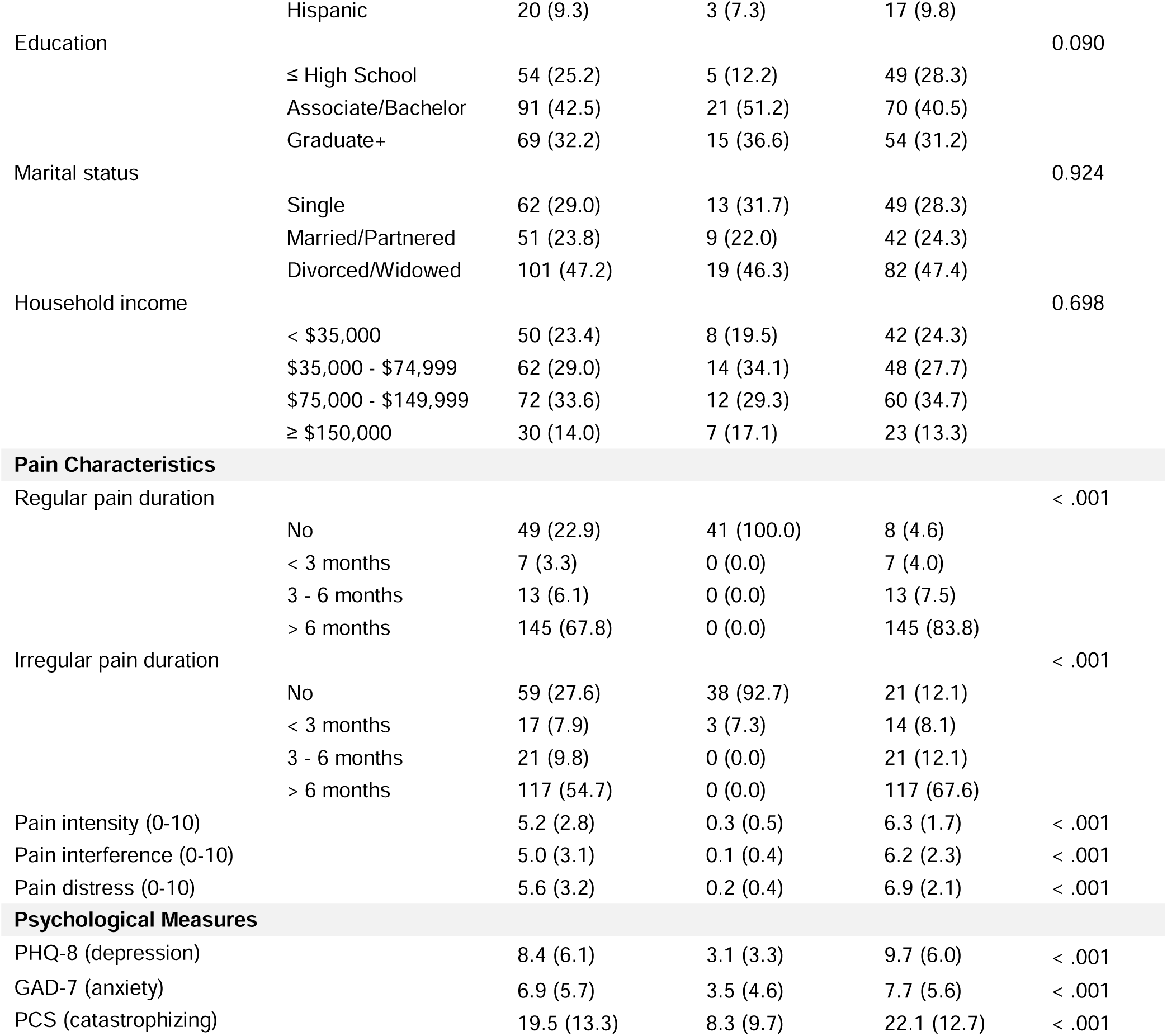
Baseline demographic and clinical characteristics by pain group. Data are presented as mean ± SD for continuous variables and n (%) for categorical variables. The Pain group includes both Acute and Chronic pain participants. Group comparisons used the Wilcoxon Rank-Sum test for continuous variables and Fisher’s exact test used Monte Carlo simulation (B=10,000 replicates) for categorical variables, to accommodate sparse cells in larger contingency tables. Abbreviations: PHQ-8, Patient Health Questionnaire-8 (depression); GAD-7, Generalized Anxiety Disorder-7; PCS, Pain Catastrophizing Scale.

### The Soma App and EMA Protocol

Pain data were collected through the Soma smartphone app (“SOMA Pain Manager”; iOS and Android), an experience sampling platform developed by the Psychiatry, Embodiment, and Computation Lab at Brown University with engineering support from the Center for Computation and Visualization (**Figure 2**) (Gunsilius et al. 2024). The app delivered three types of daily EMA-style assessments (“check-ins”), each designed to capture a different temporal dimension of the pain experience:

**Figure 2.**
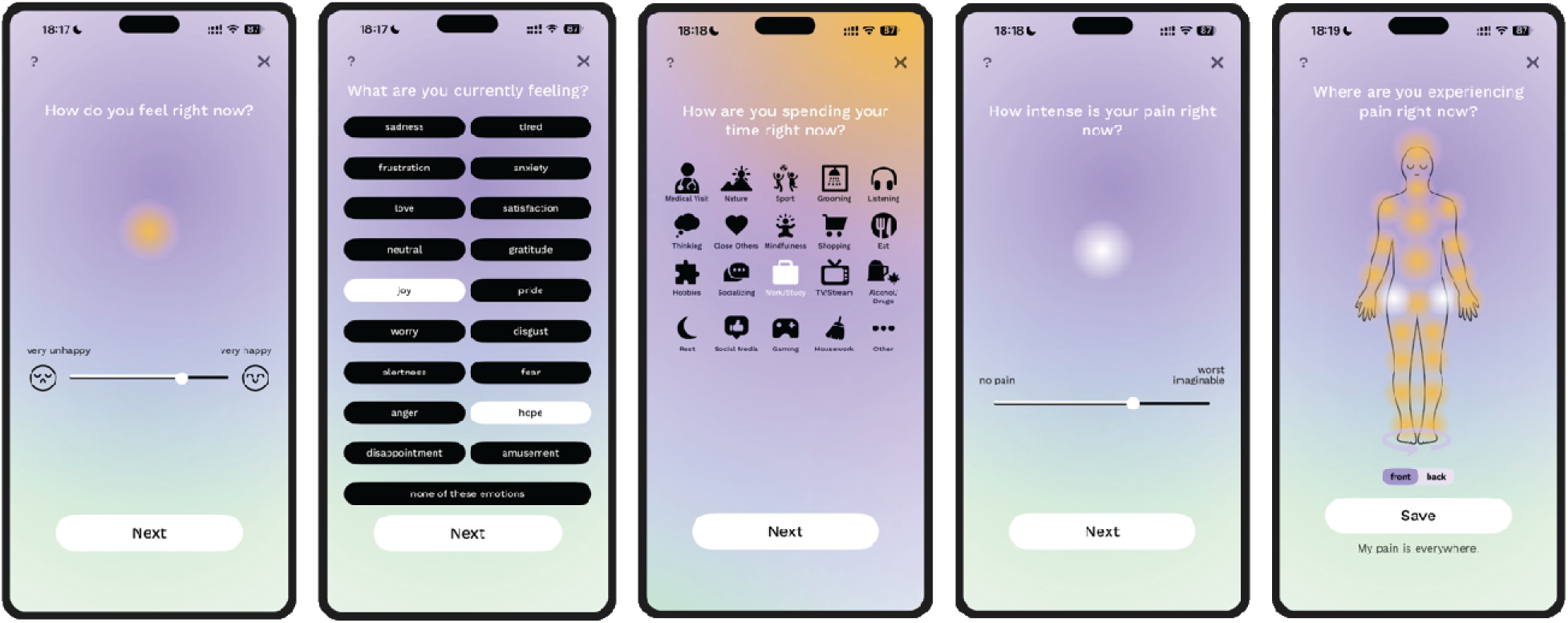
Screenshots from a momentary check-in in the Soma Pain Manager app. Four representative screens are shown from left to right: (1) pain intensity rating on a continuous visual analog scale anchored at “no pain” and “worst imaginable”; (2) current activity selection from 20 predefined categories; (3) interactive body map allowing participants to indicate current pain locations by tapping anatomical regions (front and back views, 46 selectable regions); (4) confirmation screen summarizing the selected pain locations before submission. The full momentary check-in, which also included ratings of pain unpleasantness, mood, and a discrete emotion selection (not shown), took approximately 30 seconds to complete.

#### Momentary check-ins

served as classical ecological momentary assessments. Participants received one pseudo-randomly timed push notification per day within a fixed daytime window (8:00 AM to 6:00 PM local time), which could be snoozed for up to 60 minutes. Each momentary check-in captured current pain intensity, unpleasantness, and interference on visual analog scales; current mood; a discrete emotion selection; current activities; and pain location on an interactive 46-point body map. Participants were instructed and compensated to complete one random check-in a day. Completion took approximately 30 seconds.

#### Voluntary check-ins

contained the same items as momentary check-ins but were entirely self-initiated, allowing participants to log pain at any point in time, i.e. during clinically meaningful moments such as flare-ups or sudden changes in symptoms. Unlike scheduled and prompted assessments, which constitute the traditional core of EMA protocols, self-initiated check-ins are a nontraditional EMA feature that aligns more closely with remote symptom monitoring and digital phenotyping paradigms. They were incorporated into Soma based on iterative feedback from early users, who emphasized the importance of capturing acute symptom changes outside of fixed assessment windows (Gunsilius et al. 2024).These check-ins were uncompensated, carried no prompts or reminders, and were available at any time. Data were recorded identically to momentary check-ins.

#### Evening check-ins

(Evening Routine) were available during a fixed 5-hour time window (6:00 PM to 11:00 PM local time). Unlike momentary assessments, which sample symptoms in the moment, the evening routine was designed to capture content that requires a full-day perspective: retrospective summaries of the day’s overall pain, mood, and activities; items on pharmacologic and nonpharmacologic treatments used during the day; and prospective predictions of pain, mood, and activities for the following day. The fixed evening window was chosen deliberately for a long-duration protocol. A predictable, recurring assessment time allows participants to anchor the check-in within an existing daily routine, reducing the cognitive load of remembering when to respond and likely improving sustainability over months of daily self-monitoring relative to assessments that arrive at unpredictable times. Participants were instructed and compensated to complete one evening check-in a day. Completion took approximately 3 minutes.

#### Notifications and re-engagement

Participants received daily push notifications for each scheduled check-in type. During an initial phase of the study, up to three push notifications a day were sent for momentary check-ins; this was subsequently reduced to one to reduce participant burden (Date: February 8, 2024). One push notification per day was consistently sent for the evening routine. Push notification delivery is a client-side process dependent on individual device settings, operating system version, and background app permissions; intermittent delivery failures cannot be identified from server-side records and can only be inferred from participant-reported technical difficulties. Periods of apparent non-compliance may therefore reflect undelivered prompts rather than disengagement; a limitation acknowledged in the analysis of compliance rates. When participants fail to complete both required check-ins (momentary and evening) for three calendar days in a row, automated reminder emails were sent out every day until they complete either one momentary or evening check-ins. Participants were never excluded for periods of non-use.

#### Compensation

Daily compensation was $0.50 per completed day: $0.30 for the evening check-in and $0.20 for the momentary check-in, with only the first completion of each type counting regardless of additional responses. Voluntary check-ins were not compensated. A monthly bonus of $5.00 was paid for each calendar month in which participants completed both scheduled check-in types on at least 23 of 30 days. Maximum total compensation for this part of the study, including questionnaire payments and a study-completion bonus, was $135 per participant, paid as electronic gift cards.

#### Personal trends and feedback

In addition to data collection, the app provided participants with a personal trends page that visualized their pain intensity, mood, and other tracked variables over time. This feature allowed participants to observe patterns and fluctuations in their own data across days and weeks, providing a form of ongoing personal feedback that was available but not required. While the trends page was not a study outcome, it may have contributed to sustained engagement by making the act of self-monitoring tangibly informative, particularly for participants with pain who could observe how their symptoms related to daily activities, treatments, and mood over time.

### Baseline Assessments

At enrollment (T0), a variety of baseline questionnaires were assessed which are beyond the scope of this paper and will be reported elsewhere. The key metrics assessed and reported here include: Demographic characteristics including age, sex assigned at birth, gender identity, race, ethnicity, education, marital status, and household income; Pain characteristics drawn from the eligibility screening including current regular and irregular pain status, pain duration, and single-item numerical rating scales (0–10) for pain intensity, pain interference, and pain-related distress; Psychological Measures including the Patient Health Questionnaire-8 (PHQ-8; (Kroenke et al. 2010)), screened for depressive symptom severity, the Generalized Anxiety Disorder Questionnaire (GAD-7) and the Pain Catastrophizing Scale (PCS). Baseline characteristics by pain group are reported in Table 1.

The exact wording and response options for each item are provided below.

#### Demographic items (T0 baseline)

Age: “What is your age (in years)?”. Sex at birth: “What sex were you assigned at birth (on your original birth certificate)?”. Gender: “How do you identify yourself?”. Race: “What would best describe you? (choose all that apply)”. Ethnicity: “What would best describe you?” (Hispanic / Not Hispanic). Education: “What is your highest completed education?”. Marital status: “What is your marital status?”. Household income: “What is your annual household income? (in USD)”. PHQ-8: eight standard PHQ items, summed to a total score (range 0–24).

#### Pain characteristics (eligibility screening)

Regular pain duration: “Are you currently experiencing constant pain on a regular basis?” (No / Yes, <3 months / Yes, 3–6 months / Yes, >6 months). Irregular pain duration: “Do you currently experience irregular pain (i.e. flare-ups of pain from time to time)?” (same response options). Pain intensity: “Over the past week, how intense has your pain been on average?” (0–10 NRS). Pain interference: “Over the past week, how much has the pain interfered with your life on average?” (0–10 NRS). Pain distress: “Over the past week, how much have you been bothered/distressed by your pain on average?” (0–10 NRS).

### Handling of Dropout and Missing Data

A subset of participants explicitly notified the study team that they wished to discontinue the protocol and drop out of the remainder of the study during the 122-day period (“withdrawal”, n=27), while others stopped responding without formal notice of dropout (“inactive”). In both cases, all enrolled participants who submitted at least one EMA response were retained in the analytic sample, and no participant was administratively removed for low engagement, formal withdrawal, or extended inactivity. This intent-to-include approach is analogous to a classical intent-to-treat analysis: every participant who began the protocol contributes to the primary feasibility estimates regardless of whether, when, or why they later disengaged.

Following from this principle, all primary analyses of engagement and compliance use the full 122-day protocol window as the denominator, unless stated otherwise, and days on which a participant did not submit an EMA response (whether due to formal withdrawal, silent disengagement, or transient non-response) are treated as missing and counted as zero. No imputation was performed. This is a deliberately conservative convention: a participant who withdrew on day 30 contributes 92 days of zero-compliance observations to the aggregate, pulling primary estimates downward. We adopted it because the central aim of the study is to characterize real-world feasibility of long-duration EMA, and an analysis that silently dropped post-withdrawal periods would overstate compliance by excluding precisely the participants whose disengagement is most informative for the field.

However, this conservative primary analysis conflates two distinct phenomena: inactivity among engaged participants vs disengagement from the protocol entirely. We therefore pre-specified a secondary analysis that recomputes compliance over each participant’s individual at-risk period (operationalized below and detailed in *At-Risk and Active Participant Definitions*). This is analogous to a per-protocol or on-treatment analysis in the clinical trial literature. The contrast between the full-window (”intent-to-include”) and at-risk-adjusted (”on-protocol”) estimates is a central methodological contribution of the paper.

Both analyses depend on a shared framework for classifying participant states over time. We defined sustained **disengagement** as the first occurrence of ≥14 consecutive days without any EMA check-in. A participant was classified as **at-risk** on a given study day or week if they had not yet met this criterion, that is, if their most recent check-in occurred on or after the start of that interval. Among at-risk participants, those who submitted at least one check-in during the interval were further classified as active. Participants who met the disengagement criterion but later resumed check-ins were retained in the dataset; their post-gap observations are reported where relevant. A small number of participants explicitly informed the research team of their withdrawal from the study; these are included within the disengaged group.

### Engagement Analysis

Per-user engagement metrics were derived from timestamped check-in records for the 214 participants in the analytic sample (Figure 1). Three check-in types were distinguished: evening (once daily by study protocol, fixed window 6:00–11:00 PM), momentary (once daily by study protocol, pseudo-random daytime prompt between 08:00 AM–06:00 PM), and voluntary (self-initiated at any time, uncompensated). For each participant, we computed a set of engagement metrics capturing the volume, temporal distribution, and composition of app use (Table 2). These include total check-ins overall and by type, the number of active days (calendar days with at least one check-in), the span between first and last check-in, temporal coverage (active days as a proportion of the 122-day window), check-in intensity on active days, and the median inter-check-in interval. To characterize individual differences in engagement, participants were classified into three tiers based on temporal coverage: high (≥75% of study days), moderate (25–74%), and low (<25%).

**Table 2.**
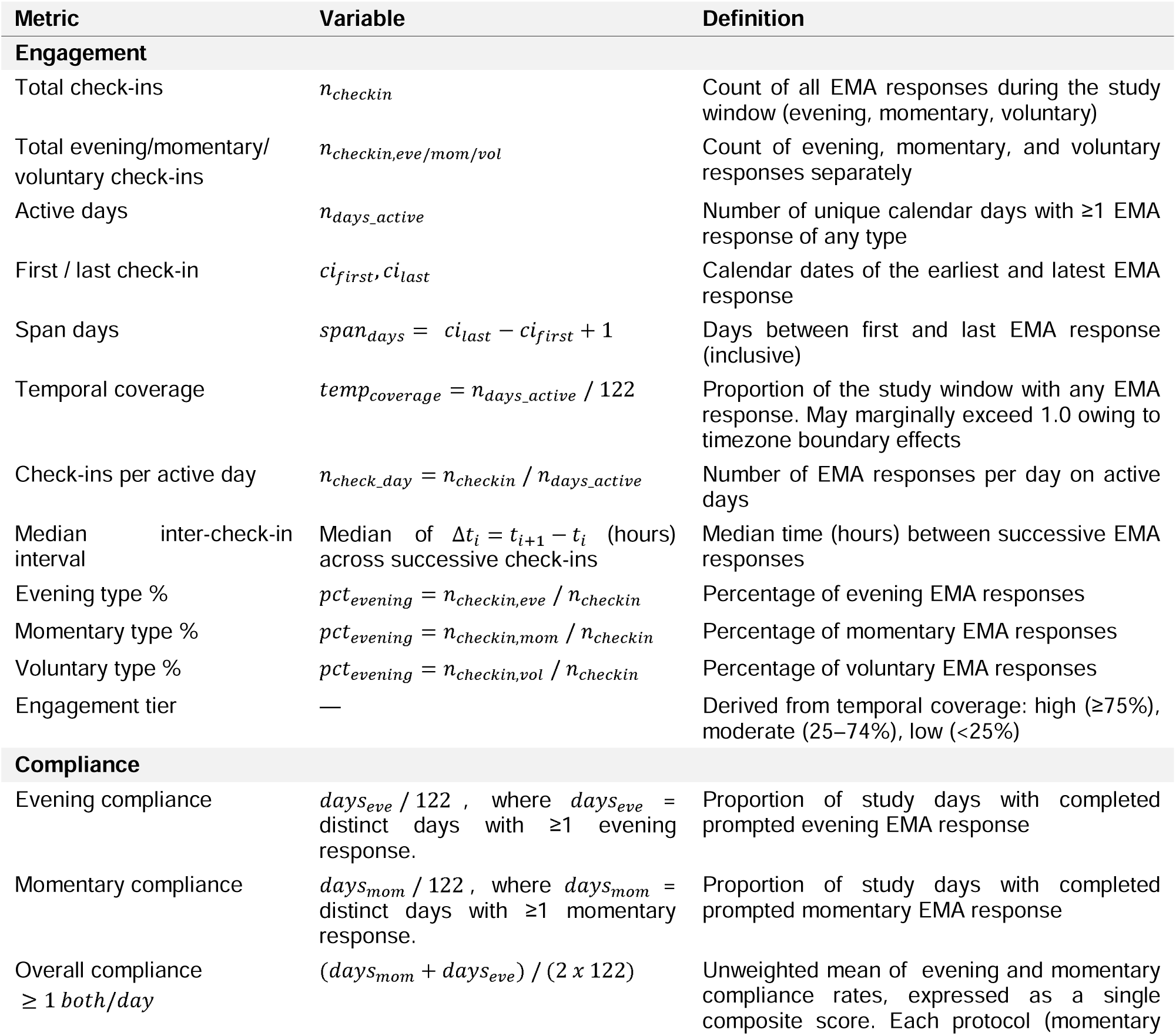
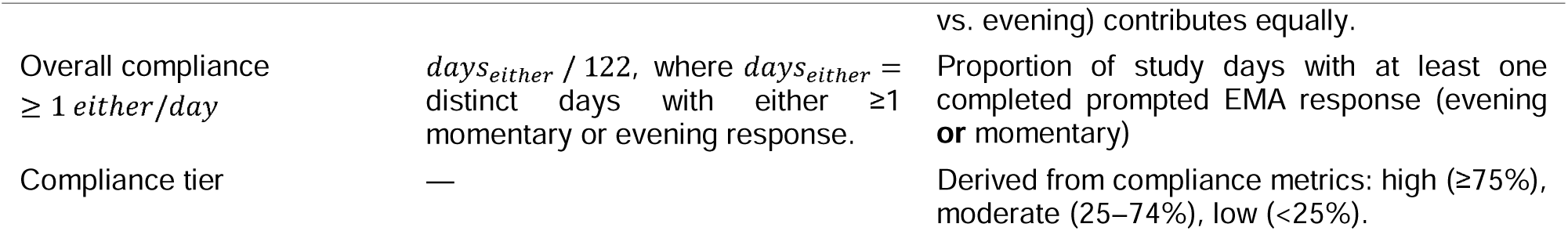
Per-user engagement and compliance metrics derived from timestamped EMA response records.

### Compliance Analysis

Protocol-based compliance was computed for prompted check-in types only (evening and momentary) and assessed separately from overall engagement. For each participant, compliance was expressed as the proportion of the 122 study days on which at least one response of the relevant type was submitted (Table 2). Two overall compliance metrics were defined: a strict criterion expressed as the unweighted mean of the evening and momentary single-protocol compliance rates (Overall compliance > 1 both/day ), and a more lenient criterion requiring at least one prompted check-in of either type (evening or momentary, Overall compliance > 1 either/day). Denominators reflect the theoretical 122-day schedule rather than prompts actually delivered, since push notification delivery cannot be tracked from server-side logs; periods of notification failure therefore appear as non-compliance. Voluntary check-ins are excluded from compliance proportions. To characterize individual differences, participants were classified into compliance tiers using the same thresholds as for engagement: high (≥75%), moderate (25–74%), and low (<25%).

### Retention Analysis

Retention was assessed using Kaplan-Meier survival analysis, with the disengagement event defined as the first occurrence of ≥14 consecutive calendar days without any EMA check-in. Disengagement data include both participants that officially withdrew from the study as well as inactive users, see section “Handling of Dropout and Missing Data” above.

A first-gap algorithm scanned all consecutive inter-check-in intervals for each participant, including the interval between the last check-in and the end of the observation window. The event was placed at the last check-in immediately preceding the first gap meeting or exceeding the threshold. Participants who never experienced a gap of this length were right-censored at their last check-in date, or at day 122 if still active at the end of the observation window.

Survival curves were estimated for the overall sample and stratified by pain group (Healthy vs. Pain), with between-group differences tested using the log-rank test. Supplementary analyses included: (i) type-specific retention curves restricted to evening, momentary, or voluntary check-ins independently, where participants with no check-ins of a given type were treated as immediate dropouts; and (ii) sensitivity analyses varying the inactivity threshold to 7 and 28 consecutive days. A Cox proportional hazards model examined baseline predictors of disengagement, entering group (pain, healthy), baseline pain intensity (0–10 NRS from eligibility screening), age, sex, and depressive symptom severity (PHQ-8) as covariates.

### Machine Learning Model for Retention Prediction

To assess whether early indicators could identify participants at risk of later disengagement, we trained an elastic net logistic regression classifier. The elastic net combined L1 (lasso) and L2 (ridge) regularization, enabling automatic feature selection. The outcome was binary disengagement, defined consistently with the retention analysis as the first occurrence of ≥14 consecutive days without a check-in at any point during the 122-day study period. To avoid circular prediction, in which the behavioral features used as predictors directly encode the outcome they are meant to forecast, we excluded 21 participants (10%) who met the disengagement criterion during the first week (survival days ≤7). This yielded an analysis sample of 192 participants (87 disengaged, 105 retained).

Nine candidate predictors comprised three categories: (i) baseline demographics (age, sex, education), (ii) baseline questionnaire scores (PHQ-8 depressive symptom severity, Pain Catastrophizing Scale total score, and pain intensity from the eligibility screen), and (iii) first-week behavioral engagement features (number of active days and total check-ins during study week 1). Sex was coded from the baseline survey; education was collapsed into three ordered levels (high school or less, associate/bachelor’s, graduate degree or higher). Anxiety (GAD-7) was excluded a priori due to high correlations with depression (PHQ-8; r = 0.74) and pain catastrophizing (PCS; r = 0.65), which would have inflated variance inflation factors (GAD-7 VIF = 2.81 with all predictors included vs. max VIF = 2.20 without). Among the retained predictors, the highest pairwise correlations were PHQ-8 × PCS (r = 0.66) and PCS × pain intensity (r = 0.55); all VIFs remained below 2.5 (range: 1.06–2.20), indicating minimal problematic multicollinearity.

The analytic sample for this analysis was partitioned into an 80-20 training-test split (Training: 80%, N = 153, Test: 20%, N = 39) stratified by disengagement status. The test set was held completely separate and used only once for final model evaluation.

To assess which predictors reliably contributed to the model, we performed a 10 × 10-fold repeated cross-validation exclusively on the training set. For each CV fold, the elastic net mixing parameter was tuned via nested 5-fold CV within the fold’s training portion, the model was fit and coefficients extracted. This yielded 100 coefficient estimates per predictor. Predictors with non-zero coefficients in >50% of folds were considered stable.

The final model was trained on the complete training set using the most frequently selected α from CV. Model discrimination was assessed using the area under the receiver operating characteristic curve (AUC) on the held-out test set, with 95% confidence intervals from 2000 bootstrap resamples.

As a secondary analysis, we used the same predictor set to predict temporal coverage (proportion of study days with ≥1 check-in) via elastic net regression, evaluated by root mean squared error (RMSE) and R².

### At-Risk and Active Participant Definitions

Two complementary denominators were used to disentangle compliance from disengagement in the weekly and adjusted analyses.

#### Weekly trajectory analyses

A participant was classified as at-risk in study week *w* if their last EMA check-in of any type occurred on or after the first day of that week. Participants whose final check-in preceded the start of week *w* were excluded from that week’s statistics. This filter prevents participants who have permanently disengaged from contributing zero-compliance observations to later weeks, which would otherwise pull weekly medians downward as dropouts accumulate, giving a misleading impression that compliance among active participants is declining. Among at-risk participants, those who completed at least one check-in during the week were further classified as *active*. Weekly compliance statistics (median, IQR) were computed over the at-risk set; engagement intensity metrics (e.g., mean check-ins per user) were computed separately over both at-risk and active subsets.

#### At-risk adjusted compliance

To compute compliance rates that exclude post-disengagement periods, each participant’s at-risk window was defined using the retention criterion established in the survival analysis: the first gap of ≥14 consecutive days without any EMA check-in. For participants who met this disengagement criterion, the at-risk window spanned from enrollment to the last check-in before the qualifying gap. For those who never met the criterion, the window spanned the full study period. Only check-ins falling within the at-risk window were included in the numerator, and the at-risk window length in days served as the denominator, replacing the fixed 122-day study duration. Participants with no check-ins (at-risk window of zero days) were excluded from this analysis.

### Statistical Analysis

Continuous variables are reported as median (IQR); the mean (SD) is additionally provided to facilitate comparison with published EMA benchmarks, which predominantly report means. Categorical variables are reported as n (%). The primary between-group comparison contrasted participants with pain (acute and chronic combined) with healthy controls using Wilcoxon rank-sum tests for continuous variables and Fisher’s exact test for categorical variables. A two-sided significance level of α = 0.05 was used throughout. Survival analyses used Kaplan-Meier estimation and Cox proportional hazards regression as described in the Retention Analysis section above.

All analyses and visualizations were performed in R version 4.4.2 (R Core Team, 2024). Key packages included survival v3.7.0 and survminer v0.5.2 for Kaplan-Meier estimation and Cox regression, ggplot2 v4.0.2 and patchwork v1.3.2 for figures, and dplyr v1.2.0 and tidyr v1.3.2 for data manipulation.

## Results

### Engagement was sustained across the full four-month protocol

Over the 122-day observation window, 214 participants (173 pain group [154 chronic pain, 19 acute pain] and 41 healthy controls) generated 26,907 EMA check-ins through the Soma app. The median participant completed 111 check-ins (IQR: 33.5–189.8; mean: 126.3, SD: 104.2, **Table 3**, **Figure 3A**) and was active on 74 of 122 available days (IQR: 24.0–107.0), roughly three out of every five study days (**Figure 3B**). On active days, participants completed a median of 1.6 check-ins (IQR: 1.3–2.0), spaced a median of 16.7 hours apart (IQR: 11.2–23.3 hours). This pattern is consistent with most participants settling into a once-daily routine, typically anchored around the evening assessment.

**Figure 3.**
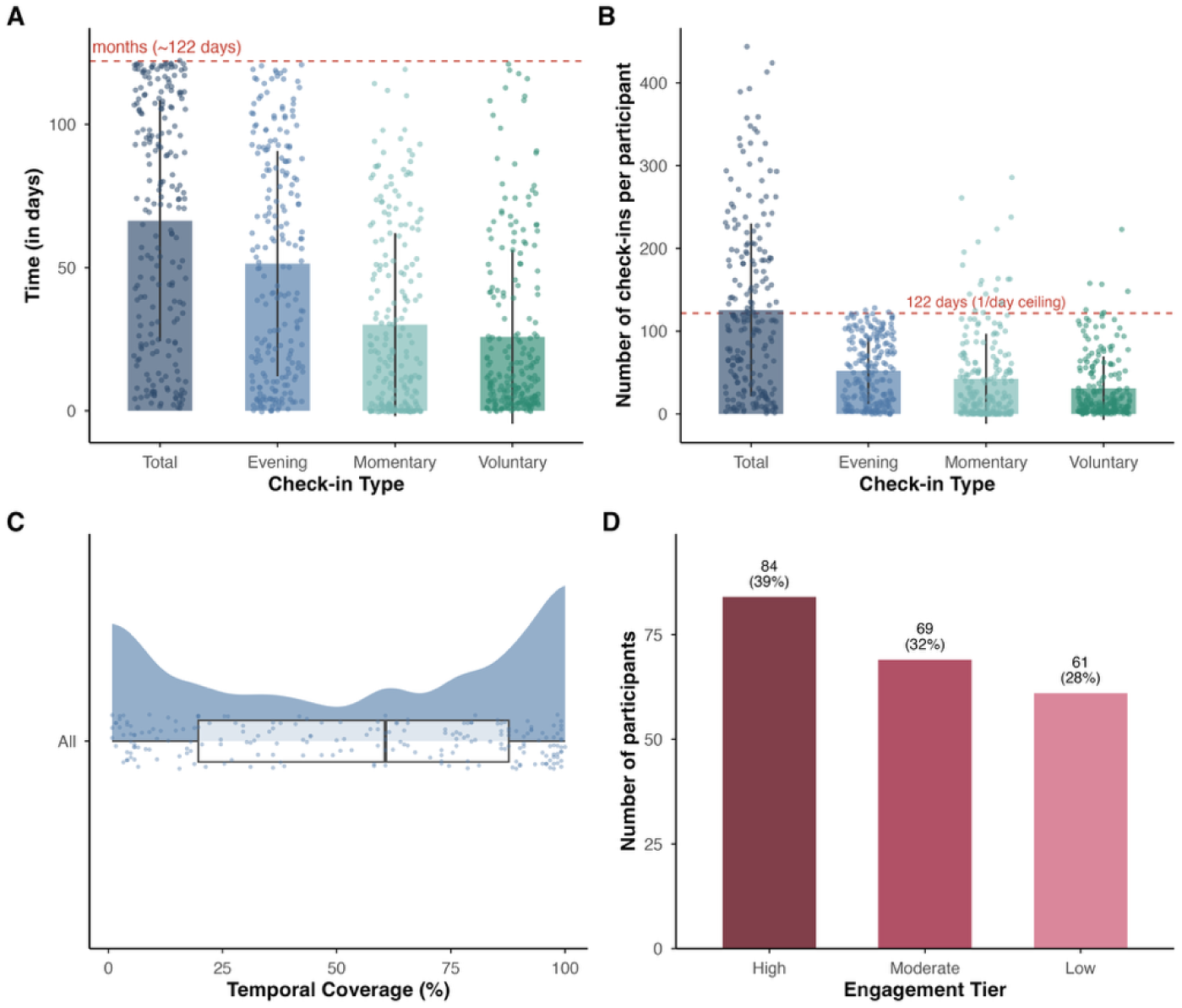
Engagement over four months across the analytic sample (N = 214). All panels are derived from per-participant metrics computed over the four-month (122-day) observation window, based on 26,907 total EMA responses. A minimum of 1 check-in total was required to be included in the analysis (n = 214). **(A)** Active days by check-in type. Bar heights represent group means (±1 SD); individual participants are shown as jittered points. The red dashed line marks the full 122-day protocol length; several participants submitted at least one EMA response on every study day. Evening check-ins account for the largest share of active days, followed by momentary and voluntary. **(B)** Total EMA responses per participant by check-in type. The dashed line indicates the theoretical ceiling of 122 responses (one per day); participants above this line submitted multiple check-in types on the same day. The evening assessment, which could only be completed once per day, shows a natural ceiling near this line. (active days / 122). The density curve and underlying boxplot (median, IQR, ±1.5 × IQR) reveal a bimodal pattern: one cluster below 25% coverage reflects participants who were relatively inactive or eventually disengaged, while a second cluster above 75% reflects sustained near-daily use across the full protocol. **(D)** Engagement tiers derived from temporal coverage: high (≥75% of study days, n = 84, 39%), moderate (25–74%, n = 69, 32%), and low (<25%, n = 61, 28%). Nearly three quarters of the sample sustained moderate-to-high engagement over four months (n=153, 71%).

**Table 3.** Summary of engagement and compliance metrics across 214 participants over the 122-day observation window. Engagement metrics: aggregate numbers of all assessments (incl. evening, momentary, voluntary EMA reports); Temporal Coverage: Proportion of the study window with any EMA response; Inter-CI interval: Median time (hours) between successive EMA responses; Engagement Tiers: Derived from temporal coverage: high (≥75%), moderate (25–74%), low (<25%). Compliance metrics: proportion of protocol-required assessments (1 evening and 1 momentary/ day EMA report); Compliance tiers: Derived from compliance metrics: high (≥75%), moderate (25–74%), low (<25%). See Table 2 for further definitions and equations. See Table 6 for additional analyses broken down by group.

| **Characteristic** |  |
| --- | --- |
| **Engagement** |  |
| Check-ins per user (median, IQR) | 111.0 (33.5–189.8) |
| Check-ins per user (mean, SD) | 125.7 (104.2) |
| Active days per user (median, IQR) (max=122 days) | 74.0 (24.0–107.0) |
| Check-ins per active day (median, IQR) | 1.6 (1.3–2.0) |
| Temporal coverage (median, IQR) | 0.6 (0.2–0.9) |
| Median inter-CI interval, hours (median, IQR) | 16.7 (11.2–23.3) |
| % Evening (median) | 41.0 |
| % Momentary (median) | 24.2 |
| % Voluntary (median) | 26.9 |
| Study span, days (median, IQR) | 111.0 (44.8–121.0) |
| Engagement tier: High ($\geq 75\%$), n (%) | 84 (39.3%) |
| Engagement tier: Moderate (25–74%), n (%) | 69 (32.2%) |
| Engagement tier: Low ($< 25\%$), n (%) | 61 (28.5%) |
| **Compliance** |  |
| Evening compliance, % (median, IQR) | 41.0 (10.0–71.9) |
| Momentary compliance, % (median, IQR) | 14.8 (1.6–42.4) |
| Overall prompted compliance, $\geq 1$ both/day, % (median, IQR) | 30.5 (8.2–54.4) |
| Overall prompted compliance, $\geq 1$ either/day, % (median, IQR) | 50.0 (15.0–82.0) |
| Compliance tier – $\geq 1$ either/day: High ($\geq 75\%$), n (%) | 69 (32.2%) |
| Compliance tier – $\geq 1$ either/day: Moderate (25–74%), n (%) | 69 (32.2%) |
| Compliance tier – $\geq 1$ either/day: Low ($< 25\%$), n (%) | 76 (35.5%) |
| Compliance tier – $\geq 1$ both/day: High ($\geq 75\%$), n (%) | 20 (9.3%) |
| Compliance tier – $\geq 1$ both/day: Moderate (25–74%), n (%) | 96 (44.9%) |
| Compliance tier – $\geq 1$ both/day: Low ($< 25\%$), n (%) | 98 (45.8%) |
| Compliance tier – Evening: High ($\geq 75\%$), n (%) | 49 (22.9%) |
| Compliance tier – Evening: Moderate (25–74%), n (%) | 77 (36.0%) |
| Compliance tier – Evening: Low ($< 25\%$), n (%) | 88 (41.1%) |
| Compliance tier – Momentary: High ($\geq 75\%$), n (%) | 9 (4.2%) |
| Compliance tier – Momentary: Moderate (25–74%), n (%) | 75 (35.0%) |
| Compliance tier – Momentary: Low ($< 25\%$), n (%) | 130 (60.7%) |

Of the 26,907 check-ins, 11,157 (41.5%) were evening assessments, 9,096 (33.8%) momentary, and 6,654 (24.7%) voluntary. At the individual level, the median composition was 41.0% evening, 24.2% momentary, and 26.9% voluntary. That roughly one in four check-ins was voluntary, self-initiated, uncompensated, and outside protocol requirements indicates that a substantial proportion of participants actively engaged in self-monitoring beyond what the study asked of them.

Crucially, this engagement was not front-loaded around enrollment. The median span from a participant’s first to last check-in was 111 days (IQR: 44.8–121.0), 91% of the full protocol length, indicating that most participants continued using the app throughout the study rather than disengaging completely after an initial period.

Temporal coverage (the fraction of available days with at least one check-in of any kind) was bimodal rather than normally distributed (**Figure 3C**): one cluster of participants fell below 25% coverage while an equally large group exceeded 85%, and one in ten participants provided near-complete daily data (90th-percentile coverage = 97.5%). In practical terms, the sample divided into two distinct engagement profiles: consistent near-daily users who maintained the protocol as designed, and episodic users.

To quantify this heterogeneity, we classified participants into three engagement tiers based on temporal coverage (**Figure 3D**): high (active on ≥75% of study days, roughly 92 or more days), moderate (25–74%), and low (<25%). Just under 40% of participants fell into the high tier (n = 84, 39.3%), maintaining near-daily app use across the full four months. About a third were moderately engaged (n = 69, 32.2%), while the remaining 28.5% (n = 61) fell into the low tier. In other words, nearly three quarters of the sample (71.5%) sustained moderate-to-high engagement over four months and the largest single group was the most engaged rather than the least.

### Compliance across 4 months was moderate but varied by participant and check-in type

Compliance, defined here as the proportion of protocol-required assessments completed, was assessed separately from overall engagement. Where engagement captures how much participants used the app in total, including self-initiated voluntary check-ins, compliance captures how reliably they fulfilled the study’s core requirement: daily completion of one evening and one momentary assessment over four full months. Compliance was computed across all 214 participants over the full 122-day window, including those who subsequently disengaged. Participants who disengaged early therefore contribute zero-compliance days for the remainder of the protocol, pulling the aggregate rates downward. An adjusted analysis restricted to each participant’s active period is presented in the below Section “Compliance Conditional on Retention”.

Under the lenient criterion (at least one prompted check-in of either type on a given day), median compliance across the full four-month period was 50.0% (IQR: 15.0%–82.0%; **Figure 4A**). Under the more stringent criterion (completing both an evening and a momentary assessment on the same day), the median was 30.5% (IQR: 8.2%–54.4%; **Table 3**). These figures reflect adherence to a demanding schedule, two prompted assessments every day for 122 consecutive days, not a brief burst of data collection. Put differently, the typical participant completed at least one prompted assessment on roughly half of all study days and completed the full daily protocol on nearly one in three days.

**Figure 4.**
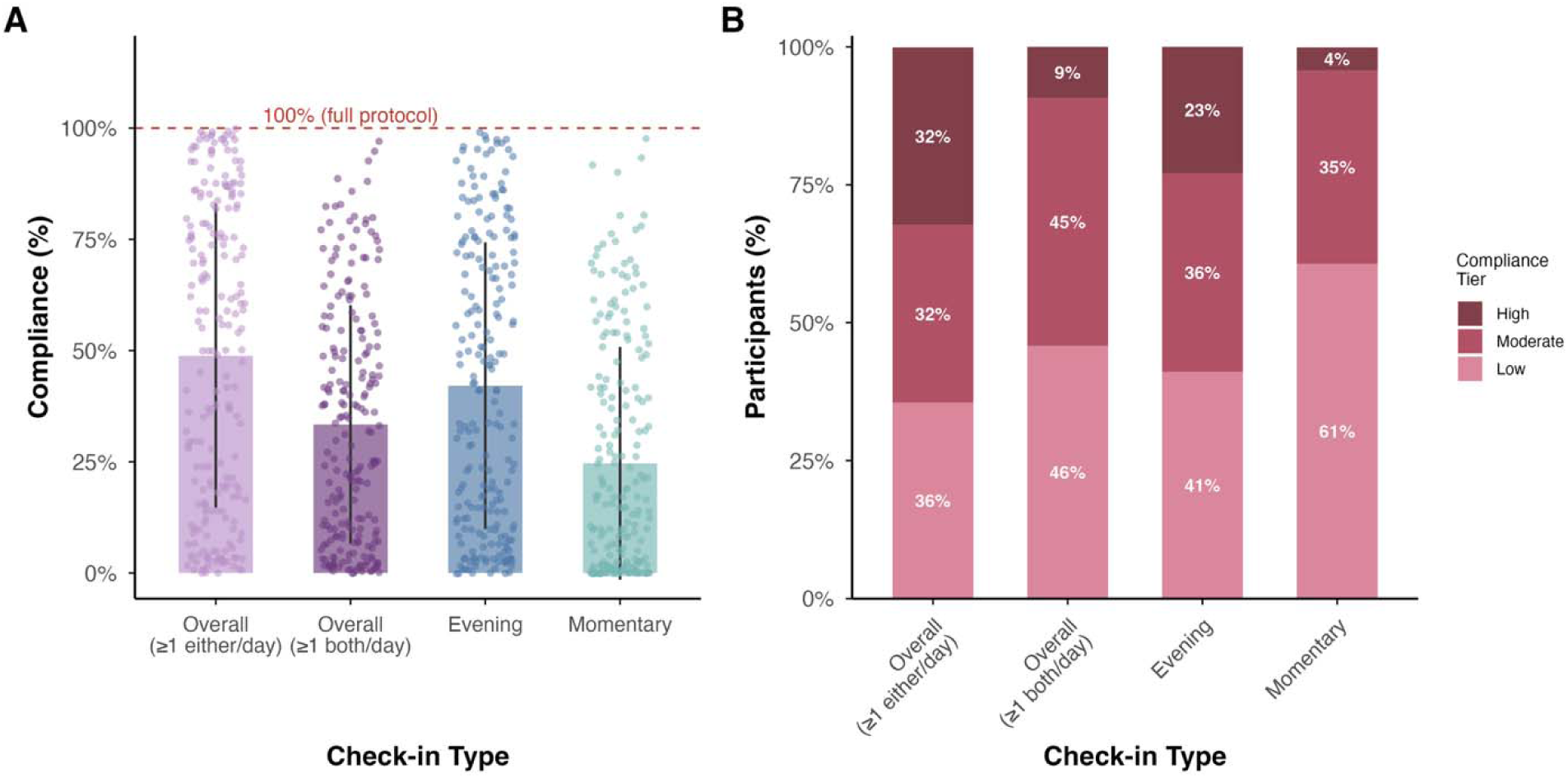
Prompted compliance across the full 4-month time window (N = 214). Compliance is defined as the proportion of the 122-day study window on which participants completed protocol-required assessments. This analysis includes all data points and participants including those who disengaged early and contributed zero check-ins for much of the observation window. All rates are computed against the theoretical maximum (one evening and/or one momentary prompt per day). Actual compliance is likely higher, as push-notification delivery failures cannot be distinguished from non-response (see text). **(A)** Per-participant compliance by criterion and check-in type. Bar heights represent group means (±1 SD); individual participants are shown as jittered points. The red dashed line marks 100% (full protocol adherence). Four metrics are shown: overall compliance under the lenient criterion (≥1 prompted check-in of either type per day), overall compliance under the strict criterion (mean of both evening and momentary check-in per day), evening compliance alone, and momentary compliance alone. The gap between evening and momentary compliance reflects the narrower response window and unpredictable timing of momentary prompts. **(B)** Distribution of participants across compliance tiers (high: ≥75%, moderate: 25–74%, low: <25%) for each criterion. Under the lenient criterion, roughly two thirds of participants achieved moderate-to-high compliance; under the strict criterion, the distribution shifted toward the lower tiers, driven primarily by low momentary compliance. Sums of slightly > 100% are due to rounding.

Compliance differed markedly by check-in type (**Figure 4A**). Evening compliance, the proportion of study days on which a participant completed the evening assessment, reached a median of 41.0% (IQR: 10.0%–71.9%). Momentary compliance, the corresponding rate for completion of at least one pseudo-random daytime prompt, was substantially lower at 14.8% (IQR: 1.6%–42.4%). This discrepancy likely reflects two compounding factors: evening assessments occurred at a predictable time and allowed a four-hour response window, whereas momentary prompts arrived unpredictably throughout the day with only a one-hour completion window, making them considerably easier to miss or dismiss. Voluntary check-ins, being self-initiated and unscheduled, are excluded from all compliance calculations.

To characterize individual differences, we classified participants into three compliance tiers based on the proportion of study days on which they met each criterion: high (≥75%), moderate (25–74%), and low (<25%) (**Figure 4B**). The distribution across tiers shifted considerably depending on which criterion was applied (**Table 3**). Under the lenient criterion, roughly two thirds of participants (64.4%) achieved moderate-to-high compliance, with the sample spreading fairly evenly across tiers (high: n = 69, 32.2%; moderate: n = 69, 32.2%; low: n = 76, 35.5%). Under the strict criterion, the distribution tilted sharply: just over half (54.2%) reached moderate-to-high compliance, and fewer than one in ten achieved the high tier (high: n = 20, 9.3%; moderate: n = 96, 44.9%; low: n = 98, 45.8%). This pattern was driven primarily by momentary assessments, where fewer than 5% of participants reached the high tier (n = 9, 4.2%), compared with nearly a quarter for evening compliance (n = 49, 22.9%).

All rates in this section should be interpreted with a methodological constraint in mind. The app infrastructure did not retain push-notification delivery logs, so we cannot determine how many prompts each participant actually received, only how many were sent. All compliance rates are therefore computed against the theoretical maximum of one evening prompt plus one momentary prompt per day across 122 days. Periods of notification failure, whether due to phone settings, operating system restrictions, or server-side errors, are indistinguishable from non-response under this denominator. The reported rates are consequently conservative estimates, and true prompt-conditional compliance is likely higher. We return to this limitation in the Discussion.

### Retention was sustained in nearly half the sample across four months

Where the preceding sections characterize how much data participants contributed and how reliably they responded to prompts across the entire 4-month period, retention addresses a more fundamental question: did participants stay active across the whole study? Specifically, we ask whether participants maintained completion of EMA assessments over the full four-month window without any major gaps, or whether they disengaged entirely at some point during the protocol. To estimate Kaplan-Meier retention curves, we defined disengagement as the first occurrence of ≥14 consecutive days without a check-in of any type. The disengagement event is placed at the last check-in preceding the qualifying gap, and participants are classified as disengaged regardless of whether they subsequently re-engaged with the app.

Of the 214 participants, 108 (50.5%) met the disengagement criterion (≥14 consecutive days without a check-in) at some point during the observation window (**Figure 5**, **Table 4**). This includes the 27 participants who formally withdrew (Average time between disengagement detection and withdrawal: median: 6 days, mean: 16.3 days, IQR: 2-19.5 days) and 81 participants who became silently disengaged. The remaining 106 (49.5%), nearly half the sample, never experienced an inactivity gap of this length across the full four-month protocol. Because the disengagement event is placed at the last check-in preceding the qualifying gap (the point at which inactivity began) rather than at the end of the gap when disengagement is confirmed, the survival curves in **Figure 5** reflect the onset of inactivity periods rather than their detection. The sensitivity analysis varying the inactivity threshold to 7 and 28 consecutive days confirmed that the overall retention pattern was robust to the disengagement definition (**Figure 5C**).

**Figure 5.**
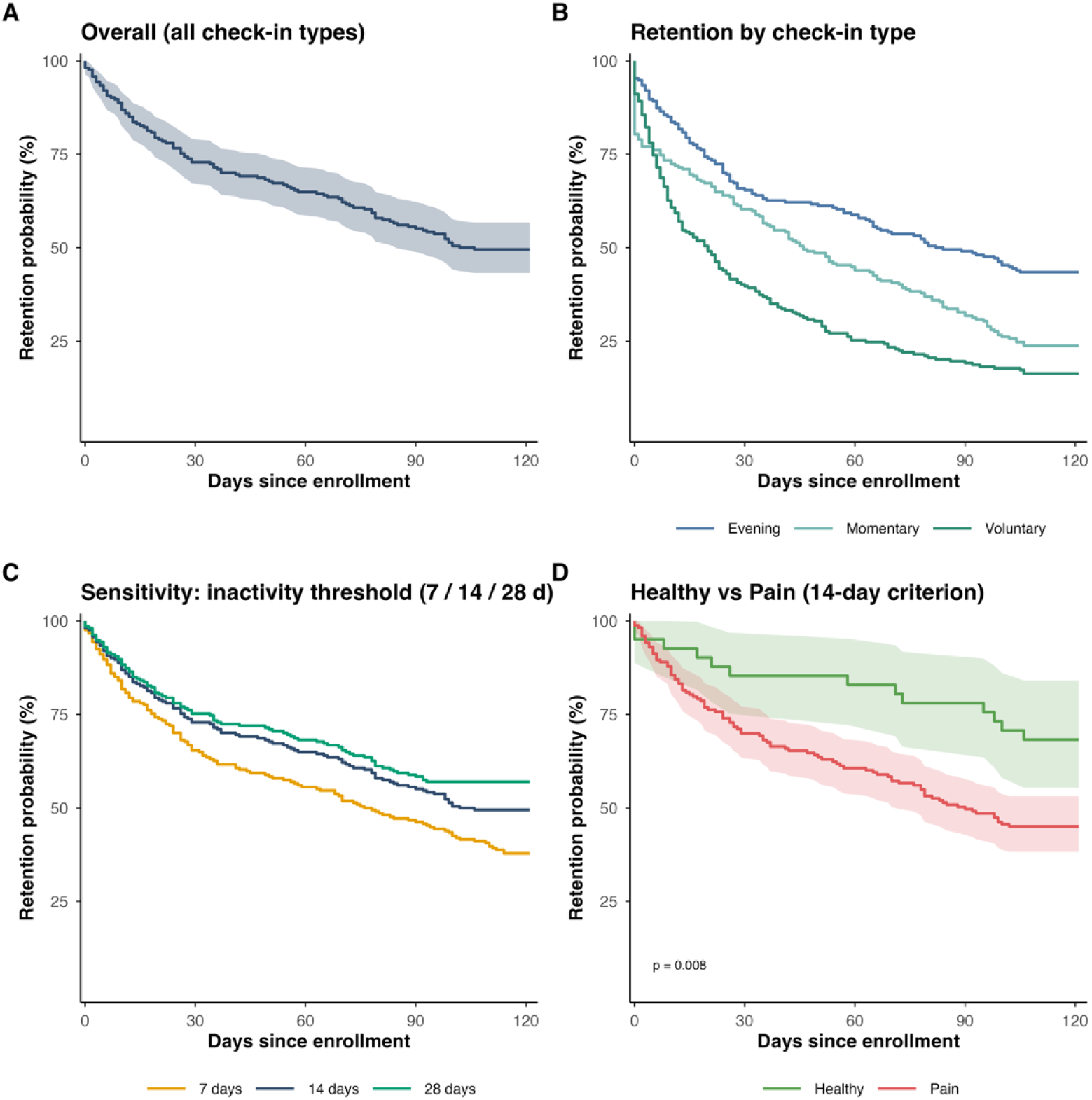
Kaplan-Meier retention curves across four months for the analytic sample (N = 214). Operational disengagement was defined as the first occurrence of ≥14 consecutive days without a check-in; the event is placed at the last check-in preceding the qualifying gap. Participants who never experienced a gap of this length were right-censored. Shaded bands represent 95% confidence intervals. **(A)** Overall retention using all check-in types combined. The curve declines most steeply in the first month, then flattens after day 30 and plateaus entirely after day 108. The final retention estimate was 49.5% (95% CI: 43.3%–56.7%). **(B)** Retention based on evening check-ins only (43.5% at day 122), momentary check-ins only (23.8% at day 122) and voluntary check-ins only (16.4% at day 122). For the type-specific panels, participants with no check-ins of that type were classified as immediate dropouts at day 0. That nearly one in six participants maintained uninterrupted voluntary engagement over four months, without prompts, reminders, or compensation, is notable. **(C)** Sensitivity analysis varying the inactivity threshold (7, 14, and 28 consecutive days). All three thresholds produced similar curve shapes, confirming the robustness of the retention estimates. **(D)** Retention by pain group (Healthy, n = 41; Pain, n = 173). Curves diverge after the first month, with Healthy controls showing significantly higher retention (log-rank p = 0.008). See **Table 4** for group-specific estimates.

**Table 4.** Kaplan-Meier retention estimates. Disengagement is defined as the first occurrence of consecutive days without a check-in meeting or exceeding the specified threshold (≥14 consecutive days without a check-in). Retention is the estimated probability of not yet having experienced a qualifying gap by day 122.

| Analysis | N | Events (%) | Median survival, days (95% CI) | Retention at day 122, % (95% CI) | Log-rank p |
| --- | --- | --- | --- | --- | --- |
| **Primary**<br>(14-day threshold) |  |  |  |  |  |
| Overall | 214 | 108 (50.5%) | 104 (85–NA) | 49.5 (43.3–56.7) | — |
| Healthy | 41 | 14 (31.7%) | Not reached | 68.3 (55.4–84.1) | 0.008 |
| Pain | 173 | 95 (54.9%) | 90 (72–NA) | 45.1 (38.2–53.1) |  |
| **By check-in type**<br>(14-day threshold) |  |  |  |  |  |
| Evening | 214 | 121 (56.5%) | 83 (65–NA) | 43.5 (37.3–50.6) | 0.010 |
| Momentary | 214 | 163 (76.2%) | 46 (36–65) | 23.8 (18.8–30.3) | 0.955 |
| Voluntary | 214 | 179 (83.6%) | 20 (13–26) | 16.4 (12.1–22.1) | 0.776 |
| **Sensitivity**<br>(threshold variation) |  |  |  |  |  |
| 7-day threshold | 214 | 133 (62.1%) | 77 (58–100) | 37.9 (31.9–44.9) | 0.024 |
| 14-day threshold | 214 | 108 (50.5%) | 104 (85–NA) | 49.5 (43.3–56.7) | 0.008 |
| 28-day threshold | 214 | 92 (43.0%) | Not reached | 57.0 (50.7–64.0) | 0.005 |

The Kaplan-Meier retention estimate declined most steeply in the first month, falling from 98.1% at day 0 to 73.8% at day 28 (**Figure 5A**). This early attrition reflects participants who checked in only a handful of times around enrollment before disengaging. After the first month, the curve flattened markedly: retention declined from 73.8% at day 28 to 49.5% at day 108, roughly 24 percentage points over 80 days, and then plateaued. The final overall retention estimate was 49.5% (95% CI: 43.3%–56.7%; **Table 4**) at day 122.

This first-gap criterion is deliberately conservative. Re-engagers who resumed check-ins after a gap of two weeks or more are classified as disengaged at the time of their initial disengagement. In practice, intermittent re-engagement after extended gaps was observed in the data. Of the 108 participants classified as operationally disengaged, 16 (14.8%) submitted at least one EMA check-in after their qualifying disengagement date, indicating that they re-engaged with the app following the initial period of inactivity. Accordingly, the retention analysis should be read as a lower bound on sustained participation, not as an indicator of permanent attrition.

When each check-in type was analyzed independently (**Figure 5B**), evening assessments showed the most durable retention profile (43.5% at day 122), consistent with their predictable timing and four-hour response window. Momentary check-ins declined more steeply, reaching 23.8% at day 122, mirroring the lower momentary compliance rates reported above. Voluntary check-ins showed the steepest attrition (16.4% at day 122), which is expected given that these were entirely self-initiated: participants received no prompts, no reminders, and no compensation for completing them. That nearly one in six participants nonetheless maintained voluntary check-ins without a two-week lapse over four months, despite having no external incentive to do so, underscores a degree of self-motivated engagement in the sample.

Retention differed significantly by pain group (**Figure 5D**). At day 122, the Kaplan-Meier estimate was 68.3% (95% CI: 55.4%–84.1%) for Healthy controls and 45.1% (95% CI: 38.2%–53.1%) for Pain participants (log-rank p = 0.008). Median survival was not reached for the Healthy group, indicating that the majority remained active throughout the protocol. For the Pain group, median survival was 90 days, meaning that half of Pain participants experienced their first two-week gap within the first three months.

In a Cox proportional hazards model with pain group, age, and sex as covariates, pain group was a significant predictor: participants with pain had approximately twice the instantaneous disengagement hazard of Healthy controls (HR = 2.26, 95% CI: 1.260–4.045, p = 0.006). Age was independently protective, with each additional year associated with a 1.8% reduction in dropout risk (HR = 0.982, 95% CI: 0.970–0.994, p = 0.003). Sex was not significant (HR = 1.120, 95% CI: 0.681–1.842, p = 0.654). The group difference was robust across all three inactivity thresholds (7-day: p = 0.024; 14-day: p = 0.008; 28-day: p = 0.005; **Table 4**).

### Can Early Engagement Patterns Predict Who Will Disengage?

The retention analysis identified pain status and age as significant predictors of disengagement at the group level. A natural follow-up question is whether individual disengagement risk can be estimated from information available early in the study, before inactivity occurs. If so, targeted re-engagement efforts could be directed toward participants most likely to disengage. To test this, we trained a machine learning model (elastic net logistic regression classifier) using baseline characteristics and first-week behavioral data to predict subsequent disengagement (see Methods).

The classifier achieved a CV-AUC of 0.70 (N = 153, IQR: 0.60 – 0.79) on the training set (internal validation). When evaluated against the held-out test set, the model showed modest discrimination (n = 39; AUC = 0.57, 95% CI: 0.37 – 0.75, CI includes chance .50), consistent with the cross-validated estimate of .70 (IQR 0.60-0.79) but with substantial uncertainty given the small test sample (**Figure 6A**).

**Figure 6.**
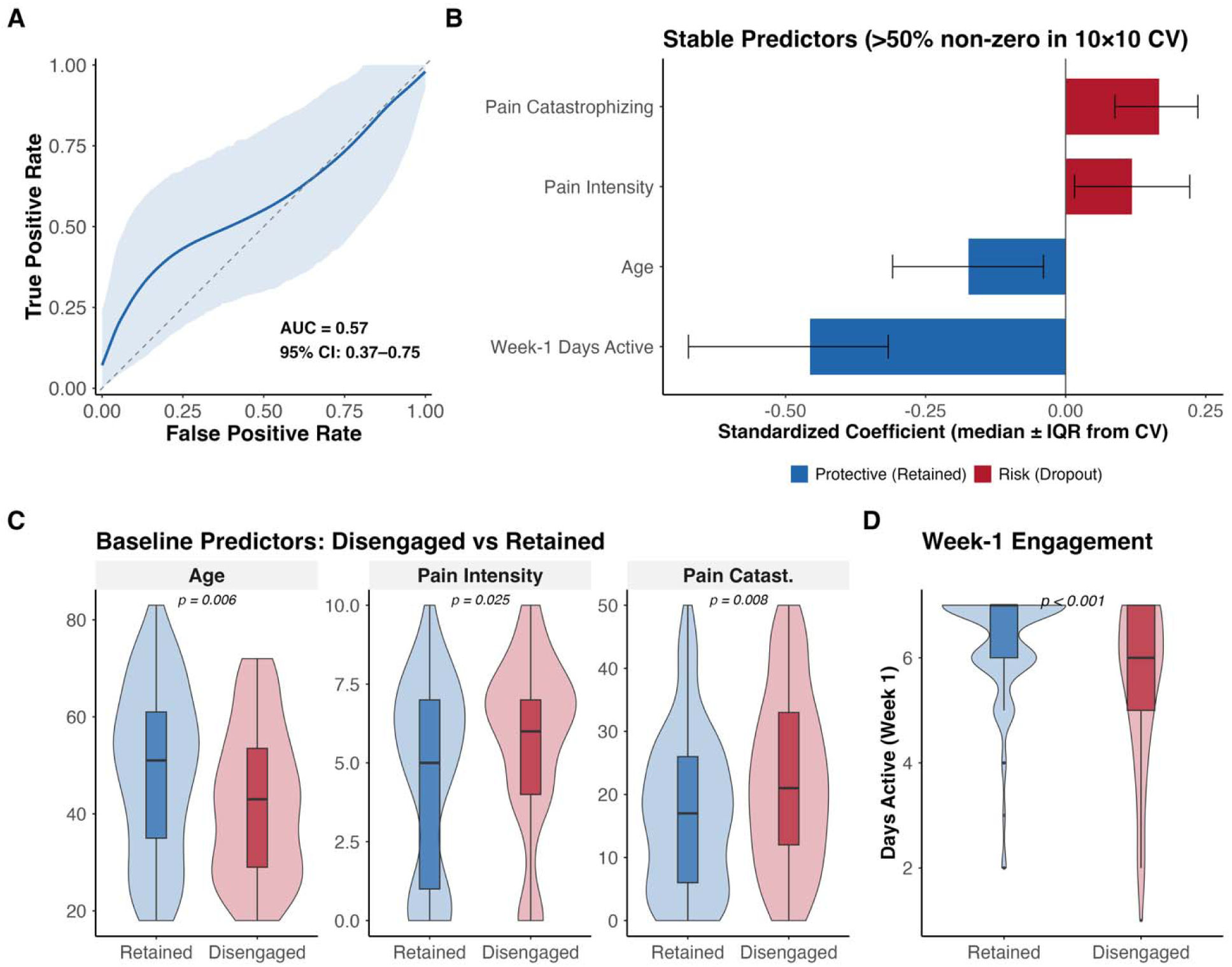
Machine learning prediction of dropout from baseline and first-week engagement data (N = 192). Participants who disengaged during the first week (n = 21) were excluded to avoid circular prediction. The model is an elastic net logistic regression trained on 80% of the data and evaluated on a held-out test set (n = 39). **(A)** Receiver operating characteristic curve. The solid line shows classifier performance on the held-out test set (AUC = 0.57); the shaded band represents the 95% bootstrap confidence interval (2,000 resamples); the dashed diagonal indicates chance-level discrimination. **(B)** Standardized elastic net coefficients for the four stable predictors (>50% non-zero across 10×10 repeated cross-validation). Red bars indicate risk factors for disengagement (positive coefficients); blue bars indicate protective factors associated with retention (negative coefficients). Error bars show the interquartile range across CV folds. **(C)** Distribution of baseline predictors (age, pain intensity, pain catastrophizing) for participants who were retained versus those who disengaged. Violin plots with embedded box plots; p-values from Wilcoxon rank-sum tests. **(D)** Distribution of first-week active day for retained versus disengaged participants. P-values from Wilcoxon rank-sum tests.

Four of the nine candidate predictors were stably selected by the elastic net, retaining non-zero coefficients in >50% of the 100 model fits of the 10x10 repeated cross-validation: Week-1 days active (100% of folds), pain catastrophizing (96% of folds), age (82% of folds), and pain intensity (77% of folds) **(Figure 6B).** The same four predictors also difference significantly between disengaged and retained participants in Wilcoxon rank-sum Tests (**Figure 6C-D**; week-1 days active p<.0001; pain catastrophizing p=.008; age p=.006; pain intensity p=.025). Of these four stable predictors, only Week-1 Days Active and Pain Catastrophizing retained non-zero coefficients at the final regularization parameter; the model’s discriminative signal is therefore driven by Week-1 engagement. The remaining predictors, sex, education, depression and week-1 check-ins – were not reliable. The protective effect of age is consistent with the Cox model reported above. That first-week days active carried substantial weight confirms what the Kaplan-Meier curves suggested: early engagement behavior is informative about long-term retention. Participants who disengaged were younger, reported higher pain intensity and pain catastrophizing, and showed lower first-week engagement.

For the secondary analysis predicting temporal coverage, the elastic net explained 29% of variance (R² = 0.29, RMSE = 0.26), improving over the null model (RMSE = 0.31). This suggests that the baseline predictors capture a moderate fraction of the factors driving engagement intensity.

### Compliance Conditional on Retention

The prompted compliance rates reported above use the full 122-day window as the denominator for all participants, including those who disengaged early. This is the appropriate conservative metric, but it conflates two distinct phenomena: non-compliance (failing to respond to a prompt while still active) and non-participation (becoming consistently inactive). To disentangle these, we recalculated all compliance metrics using each participant’s individual at-risk period as the denominator: the number of days from enrollment to their disengagement event, or to the end of the observation window for those who never disengaged. Check-ins occurring after a participant’s disengagement date were excluded.

The adjustment substantially changed the picture (**Table 5**, **Figure 7**). Under the lenient criterion (≥1 prompted check-in of either type per day), median at-risk adjusted compliance rose from 50.0% to 70.5% (IQR: 45.3%–87.5%). Under the strict criterion, the median increased from 30.5% to 41.9% (IQR: 25.1%–57.8%). Evening compliance rose from 41.0% to 58.6% (IQR: 33.4%–79.9%), and momentary compliance from 14.8% to 21.3% (IQR: 1.6%–52.4%). The shift was equally visible in the tier distributions (**Figure 7B**). Under the lenient criterion, the proportion of participants classified as moderate-to-high compliant rose from 64.5% to 91%; under the strict criterion, from 54.2% to 76%. In other words, when the denominator reflects the period during which participants were actually active and engaged with the protocol, the typical participant completed at least one prompted assessment on seven out of every ten days, and the vast majority sustained moderate-to-high compliance throughout their active period.

**Figure 7.**
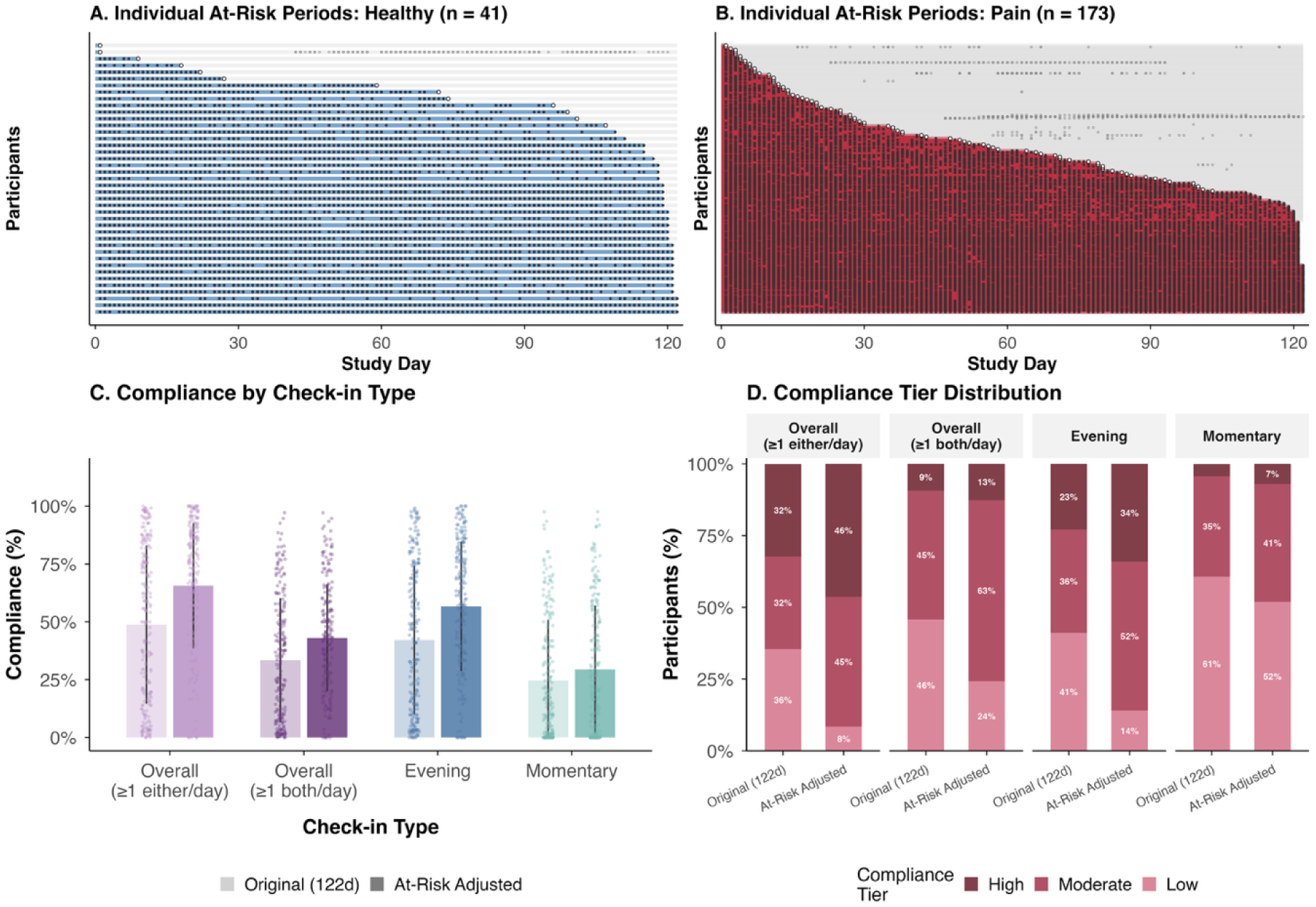
Individual activity and Prompted compliance adjusted for retention (n = 214). **(A & B)** Participant’s individual at-risk period (colored bars) and daily check-in activity (dots) over the 122-day period, sorted by at-risk duration within group (left: healthy; right: pain). Colored segments indicate the at-risk period from enrollment to the participant’s disengagement event (for those who met the ≥14-day inactivity criterion) or to day 122 (for those who did not). Gray segments indicate post-disengagement days. Dark dots represent check-ins within the at-risk window; hollow circles indicate re-engagement check-ins after disengagement. Open circles mark the time of detected disengagement. **(C)** Per-participant compliance by criterion and check-in type. Light bars: original rates (122-day denominator); dark bars: at-risk adjusted rates. Individual participants are shown as jittered points. The upward shift across all four metrics reflects the removal of post-dropout zero-compliance days from the denominator. **(D)** Compliance tier distribution (high: ≥75%, moderate: 25–74%, low: <25%) before and after at-risk adjustment. Under the lenient criterion, the proportion of participants in the high tier nearly doubles after adjustment; under the strict criterion, the share in the low tier drops substantially.

**Table 5.**
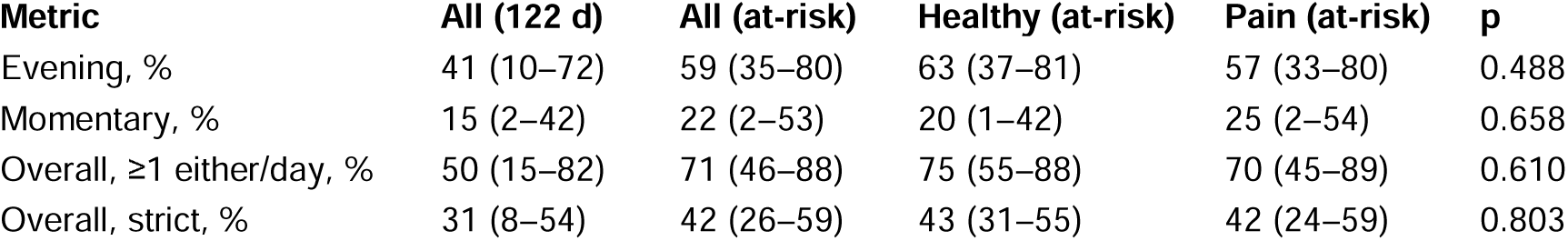
Prompted compliance computed over the full 122-day window versus each participant’s individual at-risk period. Values are median (IQR) expressed as percentages. The at-risk period reflects the number of days from enrollment to the participant’s last check-in before their first ≥14-day inactivity gap (for dropouts) or to study completion (for completers). P-values compare Healthy vs. Pain groups on the at-risk adjusted metrics using Wilcoxon rank-sum tests.

| Metric | All (122 d) | All (at-risk) | Healthy (at-risk) | Pain (at-risk) | p |
| --- | --- | --- | --- | --- | --- |
| Evening, % | 41 (10–72) | 59 (35–80) | 63 (37–81) | 57 (33–80) | 0.488 |
| Momentary, % | 15 (2–42) | 22 (2–53) | 20 (1–42) | 25 (2–54) | 0.658 |
| Overall, $\geq 1$ either/day, % | 50 (15–82) | 71 (46–88) | 75 (55–88) | 70 (45–89) | 0.610 |
| Overall, strict, % | 31 (8–54) | 42 (26–59) | 43 (31–55) | 42 (24–59) | 0.803 |

### Compliance and Engagement Did Not Decline Over Time Among Active Participants

The at-risk adjusted compliance rates in the preceding section removed the diluting effect of post-disengagement zeros from the aggregate figures, demonstrating that active participants were substantially more compliant than the unadjusted rates suggested. But an aggregate correction, even over the at-risk period, cannot reveal whether compliance was stable throughout or whether it started high and gradually eroded. If the latter, even the adjusted rates would overstate what researchers can expect from the final months of a long protocol. To address this, we examined weekly engagement and compliance trajectories across 17 study weeks (**Figure 8**).

**Figure 8.**
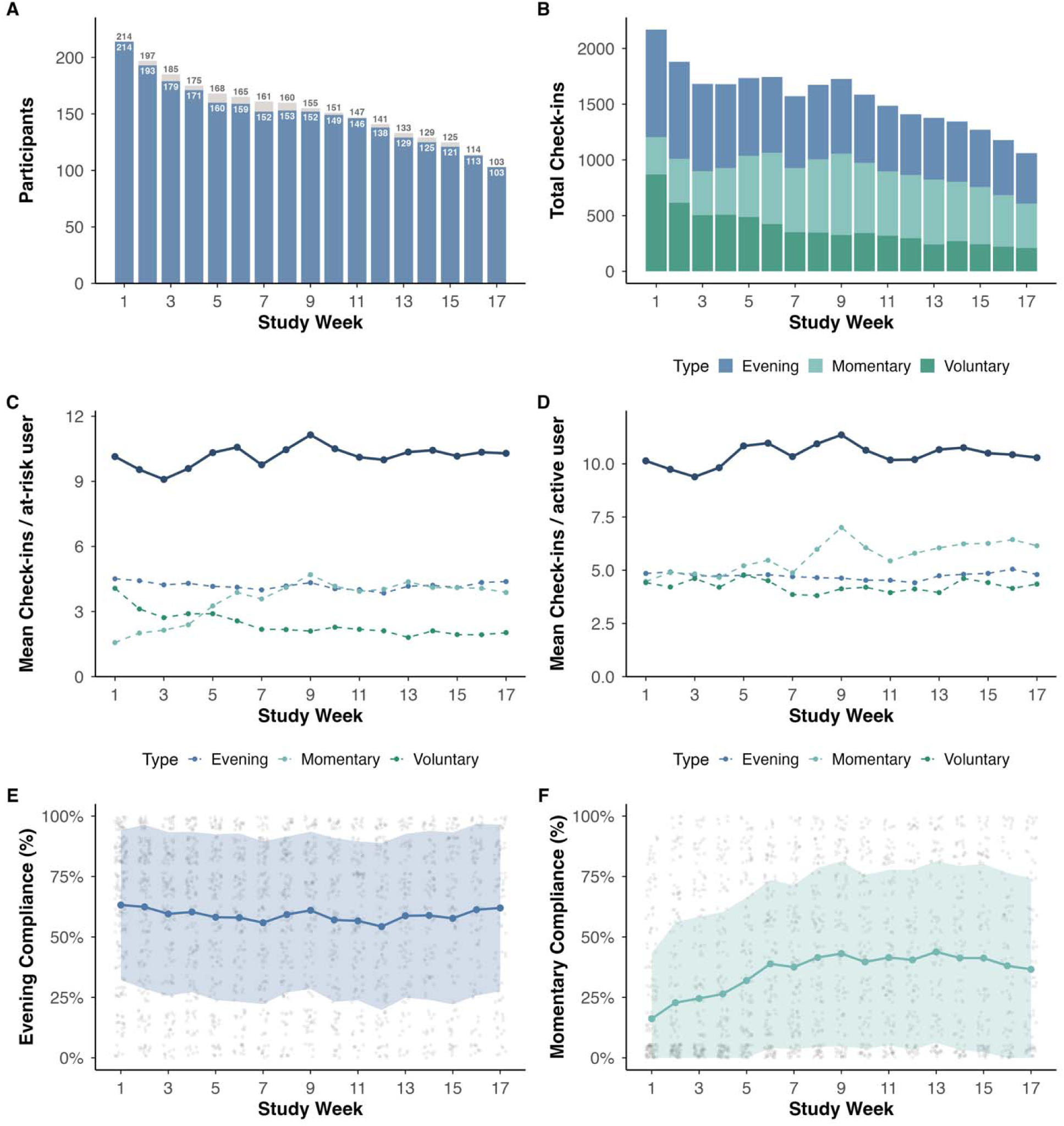
Weekly engagement and compliance trajectories across 4 months (N = 214). Study weeks are indexed from each participant’s enrollment date, not calendar date; weeks 1–17 cover study days 1–119. The final three days (120–122) are excluded to avoid artefactually inflated estimates from a truncated week. A participant is at risk in week *w* if their last check-in fell on or after the start of that week; a participant is active if they were at risk and submitted ≥1 check-in during that week. **(A)** Number of participants per week. Dark bars: active participants; light bars: at-risk participants. Numbers above bars indicate counts. The gap between bars represents at-risk participants with zero check-ins that week. **(B)** Total check-in volume by type and week. The decline over time is driven by attrition among disengaged users rather than reduced output among active participants. **(C)** Mean check-ins per at-risk participant per week, by type. The denominator includes all at-risk participants, including those with zero check-ins, so this metric is pulled downward by inactive-but-not-yet-disengaged participants. **(D)** Mean check-ins per active participant per week, by type. Restricted to participants with ≥1 check-in that week. The stable trajectories indicate that engagement intensity among active participants did not decline over the study period. **(E)** Weekly evening compliance among at-risk participants. Line shows the median; shaded band spans the interquartile range; individual participant trajectories are shown as semi-transparent lines. **(F)** Weekly momentary compliance among at-risk participants, plotted as in panel E.

The number of at-risk participants declined steadily from 214 in week 1 to 103 by week 17, reflecting the cumulative disengagement documented in the retention analysis (**Figure 8A**). Yet among those still at-risk in any given week, the vast majority remained active: even in week 17, 103 of 103 at-risk participants (100%) submitted at least one check-in. The decline in total check-in volume over the study (**Figure 8B**) was therefore driven almost entirely by attrition, not by fading engagement among those who stayed active.

Two intensity metrics confirm this (**Figures 8C–D**). Mean check-ins per at-risk participant (which includes those with zero check-ins in a given week; see Methods) and mean check-ins per active participant (restricted to those who submitted ≥1 check-in that week) were both largely stable across the full four months. Active participants provided approximately 10 EMA submissions per week from week 1 through week 17, and the type-specific trajectories (evening, momentary, voluntary) showed no systematic decline.

Weekly compliance trajectories told a similar story (**Figures 8E–F**). Among at-risk participants, median evening compliance remained between approximately 57% and 71% across all 17 weeks, with no discernible downward trend. Momentary compliance, while lower in absolute terms, was similarly stable and showed an increasing trend over time. The wide interquartile bands reflect the between-person heterogeneity documented in the tier analyses, but the central tendency did not drift.

This temporal stability completes the picture begun in the preceding sections. The unadjusted compliance rates above appeared moderate because they included post-disengagement zeros. The at-risk adjusted rates showed that active participants were in fact substantially more compliant. The weekly trajectories now confirm that this higher compliance was not concentrated in the early weeks but sustained throughout the protocol. Taken together, the compliance that researchers can expect from retained active participants in a four-month EMA study is both higher than unadjusted rates suggest and stable over time.

### Pain Status Affected Retention but Not Engagement

The retention analysis established that participants with pain were significantly more likely to disengage than Healthy controls (45.1% vs 68.3% at day 122; log-rank p = 0.008; Cox HR = 2.26, p = 0.006; **Figure 5D**, **Table 4**). The present section examines whether pain status also affected the level of engagement and compliance among those who participated, regardless of how long they stayed active.

In aggregate, Healthy controls showed numerically higher engagement on several metrics (**Table 6**, **Figure 9A–B**): more active days (median 89 vs 65, p = 0.054), higher temporal coverage (73% vs 53%, p = 0.054), and higher evening compliance (56% vs 34%, p = 0.016). Overall prompted compliance under the lenient criterion was also significantly higher in Healthy controls (66% vs 44%, p = 0.029), as was the proportion of participants in the high engagement tier (49% vs 37%; Fisher’s exact p = 0.029; **Figure 9C–D**). These differences are consistent with the higher disengagement rate in the Pain group: participants who leave the study early contribute fewer active days and lower temporal coverage by definition, pulling the group-level medians downward.

**Figure 9.**
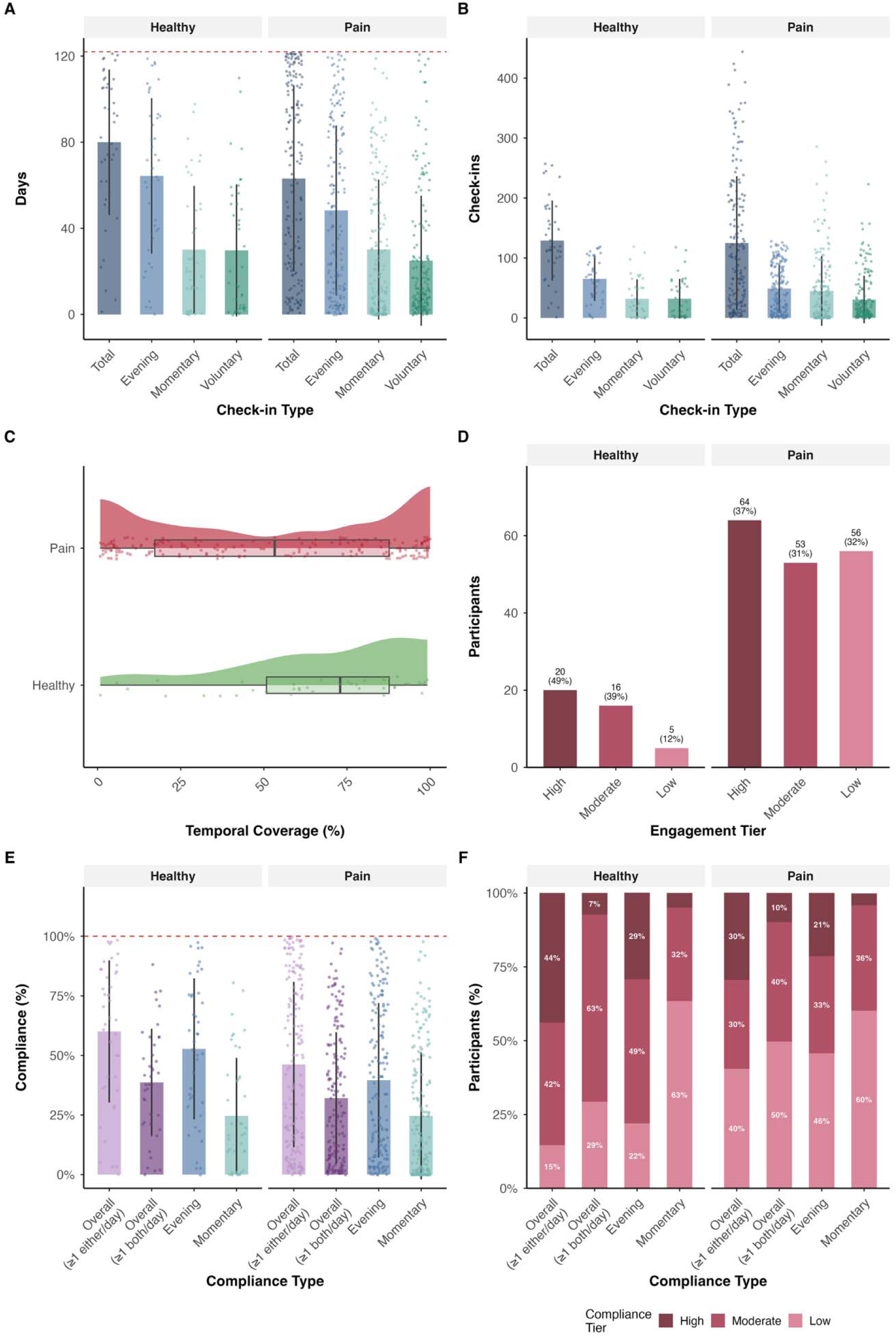
Engagement and compliance by pain group (Healthy, n = 41; Pain, n = 173). All panels are derived from per-participant metrics computed over the 122-day observation window. **(A)** Active days by check-in type and group. **(B)** Total check-ins per participant by check-in type and group. **(C)** Distribution of temporal coverage by group. Healthy controls show a unimodal distribution shifted toward higher coverage; Pain participants show a bimodal pattern with clusters at both low (<25%) and high (>75%) coverage. **(D)** Engagement tiers by group (high: ≥75%, moderate: 25–74%, low: <25% of study days). Healthy controls were significantly more likely to fall in the high tier (Fisher’s exact p = 0.029). **(E)** Per-participant compliance by criterion and group. The red dashed line marks 100% (full protocol adherence). **(F)** Compliance tier distribution by criterion and group.

**Table 6.** Engagement and compliance metrics by pain group across the full four month window (including Disengaged users). Values are median (IQR) unless otherwise noted. P-values from Wilcoxon rank-sum tests (continuous) or † Fisher’s exact test (categorical). Compliance rates are computed against the theoretical 122-day maximum.

| Metric | Healthy (n = 41) | Pain (n = 173) | p |
| --- | --- | --- | --- |
| **Engagement** |  |  |  |
| Total check-ins | 118.0 (95.0–182.0) | 98.0 (27.0–199.0) | 0.220 |
| Active days | 89.0 (62.0–107.0) | 65.0 (21.0–107.0) | 0.054 |
| Temporal coverage, % | 73.0 (50.8–87.7) | 53.3 (17.2–87.7) | 0.054 |
| Check-ins per active day | 1.6 (1.3–1.8) | 1.7 (1.3–2.0) | 0.190 |
| Inter-check-in interval, hours | 17.1 (13.7–23.4) | 16.6 (10.4–23.3) | 0.208 |
| Composition: % evening | 46.0 (35.6–57.7) | 40.0 (30.6–56.1) | 0.073 |
| Composition: % momentary | 17.6 (5.4–37.8) | 26.1 (4.3–43.9) | 0.195 |
| Composition: % voluntary | 27.0 (3.8–46.0) | 26.7 (9.2–45.6) | 0.406 |
| Engagement tier: High | 20 (48.8%) | 64 (37.0%) | 0.029† |
| Engagement tier: Moderate | 16 (39.0%) | 53 (30.6%) |  |
| Engagement tier: Low | 5 (12.2%) | 56 (32.4%) |  |
| **Compliance** |  |  |  |
| Evening compliance, % | 55.7 (30.3–75.4) | 33.6 (8.2–70.5) | 0.016 |
| Momentary compliance, % | 16.4 (0.8–41.0) | 13.9 (1.6–43.4) | 0.797 |
| Overall compliance, ≥1 either/day, % | 65.6 (36.9–85.2) | 44.3 (12.3–79.5) | 0.029 |
| Overall compliance, ≥1 both/day, % | 37.7 (21.7–54.5) | 25.0 (7.0–54.1) | 0.083 |

The at-risk adjusted compliance analysis confirmed this interpretation (Table 5). Under the full 122-day denominator, Healthy controls showed significantly higher evening compliance (56% vs 34%, W = 4405.5, p = 0.016) and lenient overall compliance (66% vs 44%, W = 4325.5, p = 0.029). Once compliance was computed over each participant’s individual at-risk period, these differences narrowed to non-significance (evening: 63% vs 57%, W = 3794, p = 0.488; lenient overall: 75% vs 70%, W = 3728.5, p = 0.611). The aggregate compliance gap between groups was driven by differential disengagement, not by differences in adherence behavior among active participants.

On days when Pain participants used the app, they used it just as intensively as Healthy controls: check-ins per active day did not differ between groups (Healthy 1.6 vs Pain 1.7, p = 0.190), nor did the inter-check-in interval (17.1 vs 16.6 hours, p = 0.298; **Table 6**). The composition of check-ins was also similar, with comparable proportions of evening, momentary, and voluntary responses in both groups (**Figure 9A–B**).

The weekly trajectory data (**Figure 10**) went further, showing that among participants who remained in the study, the Pain group became progressively more engaged than Healthy controls. In the early weeks, check-in volume and compliance were comparable between groups or slightly higher in Healthy controls. By the later weeks, this reversed. From week 15 onward, Pain participants submitted significantly more check-ins per week (week 16: Pain mean 11.6 vs Healthy 6.9, p = 0.006; week 17: Pain 11.5 vs Healthy 6.9, p = 0.014; **Figure 10C–D**). Momentary compliance showed a similar crossover: Healthy controls were initially more compliant (weeks 1–3, p < 0.03), but Pain participants surpassed them from approximately week 8 onward (p = 0.035), with the difference widening in the final months (week 17: p < 0.001; **Figure 10F**). Evening compliance, by contrast, remained comparable between groups throughout the study, with no significant difference at any week (**Figure 10E**). In short, pain status predicted whether participants disengaged, but among those who stayed active, it was associated with greater, not lesser, long-term engagement.

**Figure 10.**
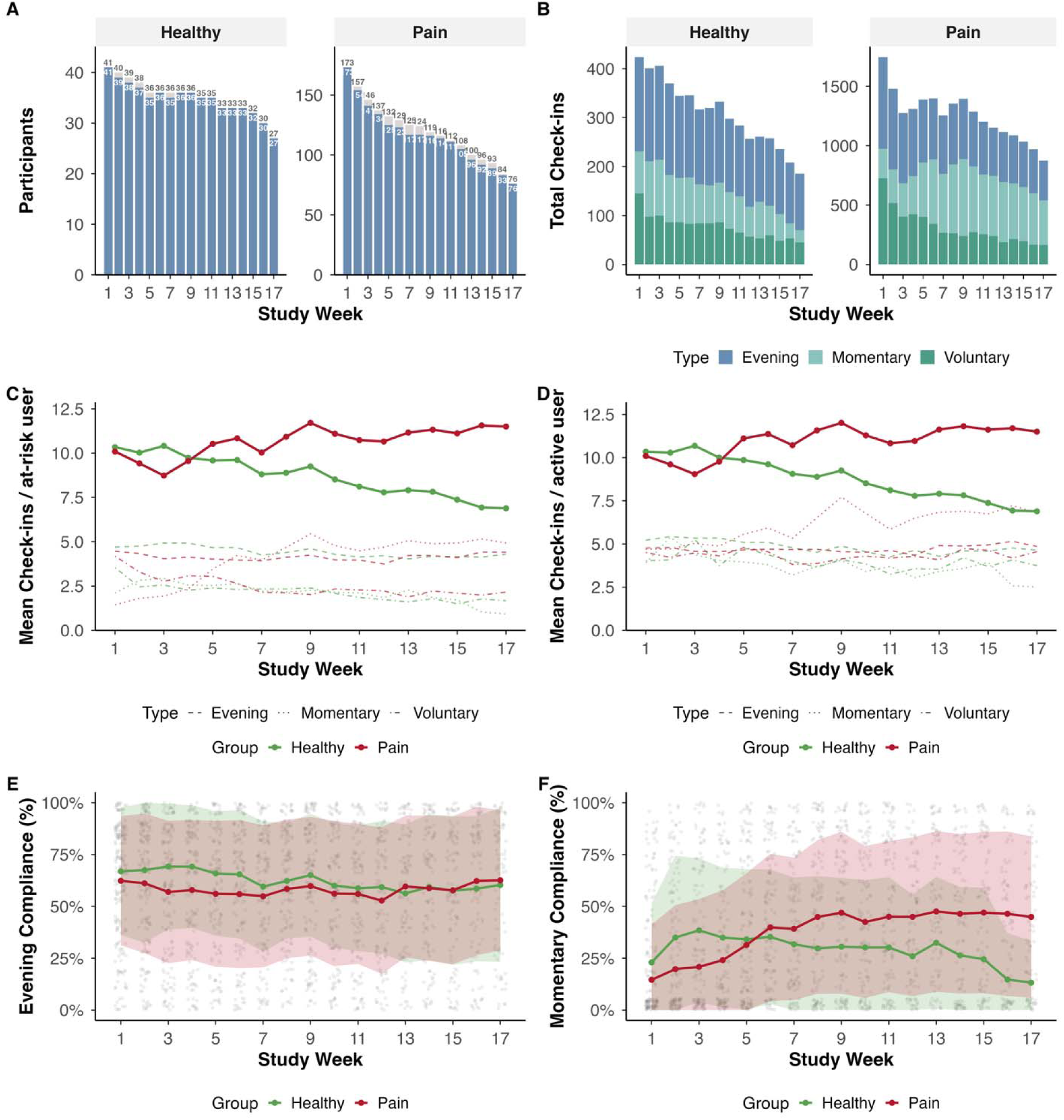
Weekly engagement and compliance trajectories by pain group (Healthy, n = 41; Pain, n = 173). Study weeks are indexed from each participant’s enrollment date; weeks 1–17 cover study days 1–119. Definitions of at-risk and active participants are as described for Figure 6. **(A)** Number of at-risk (light bars) and active (dark bars) participants per week, by group. The Pain group shows steeper attrition, consistent with the retention analysis. **(B)** Total check-in volume by type, week, and group. **(C)** Mean check-ins per at-risk participant per week, by group and type. Solid lines: total; dashed lines: type-specific. **(D)** Mean check-ins per active participant per week, by group and type. Pain participants maintained or increased their output in the later weeks while Healthy controls declined modestly. **(E)** Weekly evening compliance by group. Lines show medians; shaded bands span the interquartile range. No significant group difference at any week. **(F)** Weekly momentary compliance by group. Pain participants showed significantly higher momentary compliance from approximately week 8 onward.

## Discussion

Digital health increasingly depends on patients reporting symptoms, behaviors, and states over months or years, whether to monitor chronic disease, power decentralized trials, or detect clinically meaningful change. Yet the empirical basis for sustained self-monitoring rests almost entirely on EMA protocols lasting a week or two, leaving a basic question unanswered: can participants maintain daily reporting at the timescales clinical use actually requires? Drawing on 26,907 check-ins from 214 participants tracked continuously over 122 days, the present study offers an early detailed answer, and it turns on a distinction the field has rarely made explicit. Retention and compliance, often treated as a single construct, tell fundamentally different stories. Compliance among retained participants, regardless of group, did not decay over the four-month window. Retention, not motivational fatigue, emerged as the primary feasibility challenge. And participants living with chronic pain, despite higher dropout rates, were among the most consistently engaged users in the later months of the study.

### Compliance among active participants is stable across four months

Prompted compliance was stable across all 17 study weeks among participants who remained in the protocol. Active participants submitted approximately 10 check-ins per week from the first week to the last, and median evening compliance among at-risk participants held between 57% and 71% at every weekly timepoint, with no systematic decline in any metric. This runs counter to what the existing short-protocol literature would predict. The most comprehensive cross-field meta-analysis of EMA feasibility reports a mean compliance rate of 79% across studies lasting a median of 7 days (Wrzus and Neubauer 2023), and pooled completion rates of 75–85% have been reported in chronic pain protocols spanning 4 to 28 days (Morren et al. 2009; Ono et al. 2019), with comparable short-protocol rates documented in experience sampling studies of severe mental disorders (Vachon et al. 2019). The Ono et al. meta-analysis further estimated a decline of approximately 2.3 percentage points per week; extrapolated naively to 17 weeks, that trajectory would predict near-zero compliance by the end of the present protocol, rendering multi-month daily EMA effectively unfeasible. The present data do not support this projection.

One reason short-protocol decay rates fail to extrapolate is that they conflate two distinct phenomena that are typically treated as one. *Retention*, whether a participant remains in the protocol at all and *compliance*, how reliably retained participants respond to prompts, behave differently over long timescales, and pooling them obscures the behavior of each. The conceptual separation has been drawn before in the EMA literature (Vachon et al. 2019), but its empirical consequences emerge most clearly at the longer-duration timescales examined here, where the two come apart in ways short protocols cannot reveal. Aggregate compliance rates computed across the full 122-day window for all 214 enrolled participants were 50.4% under a lenient criterion (≥1 prompted check-in per day) and 30.8% under a strict criterion (both prompted types per day), which is well below the one-week benchmarks in absolute terms. But participants who disengaged early contribute zero-compliance days for every week after their departure, dragging the aggregate downward in a way that has nothing to do with the engagement behavior of those still active in the study.

When compliance was recomputed over each participant’s individual at-risk period (from enrollment to their disengagement event, or to day 122 for those who never disengaged), the picture changed substantially. Lenient compliance rose from 50.0% to 70.5%, evening compliance from 41.0% to 58.6%, and strict compliance from 30.5% to 41.9%. Under this adjustment, 91% of participants achieved moderate-to-high lenient compliance during their active period. These figures are considerably closer to the one-week benchmarks despite covering a window 17 times longer, and more realistically describe what researchers can expect from participants who remain in long-duration protocols under conditions resembling this one. The contrast between the unadjusted and adjusted figures illustrates a broader concern that single-number adherence summaries can mask substantial heterogeneity in how participants are actually using a digital health tool, an issue that has been raised repeatedly about engagement measurement in this literature (Torous, Michalak, and O’Brien 2020)

The contrast also has a structural parallel in clinical trial methodology. Unadjusted compliance is analogous to an intent-to-treat (ITT) estimate, computing a rate across all enrolled participants regardless of whether they remained engaged; at-risk-adjusted compliance is analogous to a per-protocol estimate, restricting the denominator to participants still actively contributing. As Hernán and Robins (2017) argue for the trial setting, the two figures answer different questions (Hernán and Robins 2017). The ITT-style aggregate addresses the representativeness of the full enrolled cohort, while the per-protocol-style figure characterizes the response behavior of those actually generating data. Informative reporting often requires both rather than either alone (Hernán and Robins 2017). The analogy is structural rather than substantive (trial estimates concern treatment effects, not descriptive engagement metrics), but the underlying logic that an aggregate computed over an enrolled cohort can misrepresent the behavior of those who actually contributed data is the same.

These observations carry direct implications for digital health protocol design. Reporting aggregate compliance across an entire enrollment cohort can substantially understate the engagement of active participants, particularly in protocols where early disengagement is concentrated, a pattern characteristic of remote digital health studies more broadly (Pratap et al. 2020) and now well documented across multiple long-duration cohorts (Zhang et al. 2023). For decentralized trials, digital biomarker validation, and remote symptom monitoring (Inan et al. 2020), where the validity of inference depends on understanding both who remains and how they engage, reporting retention and at-risk-adjusted compliance as separate quantities, rather than collapsing them into a single adherence figure, should be considered standard rather than exceptional practice.

### Disengagement is concentrated in the first month and predictable from week-one behavior and baseline characteristics

Disengagement was largely concentrated in the first month of the protocol. The retention curve fell from 98% at enrollment to 74% by week four, then flattened markedly, with no further disengagement events after day 108. This shape is consistent with the sigmoid attrition curve Eysenbach (2005) proposed for digital health interventions (Eysenbach 2005), in which a stable group of committed users remains after an initial period of steep dropout, and with the front-loaded retention patterns observed at shorter timescales in cross-study analyses of remote digital health research (Pratap et al. 2020) and app-based chronic disease interventions (Meyerowitz-Katz et al. 2020). What the present data add is that, within the four-month window, the plateau appears to represent a genuine steady state rather than a gradual slowing: both retention and compliance held constant after the first month. This inference rests on 14 days of post-plateau observation and should be confirmed in studies with longer follow-up.

Whether participants who will disengage can be identified prospectively is a separate and clinically important question. An exploratory elastic net classifier using baseline characteristics and first-week behavioral features discriminated subsequent disengagement at modest above-chance levels (AUC = 0.57), with first-week compliance and age carrying the most predictive weight. The held-out sample was small (n = 39) and this result should be treated as hypothesis-generating, but it is consistent with cross-study evidence that early engagement indicators predict long-term retention in remote digital health protocols (Pratap et al. 2020) and suggests that signals adequate to inform retention interventions may be available within the first week.

For digital health protocol design, the implication is twofold. First, retention efforts in long-duration remote studies, including onboarding support, personalized re-engagement, and adaptive incentives, should concentrate on the first month, when the retention curve is steepest and when behavioral signals of impending disengagement are already detectable. Second, the apparent steady state after the first month suggests that the marginal cost of extending a protocol from one to four months may be smaller than the short-protocol literature implies, with direct consequences for the feasibility of decentralized trials and remote chronic disease monitoring (Inan et al. 2020).

### Apparent group differences in engagement reflect differential retention

Participants with pain were retained at significantly lower rates than Healthy controls (45% vs 68% at day 122; Cox HR = 2.26, p = 0.008), and aggregate compliance metrics across the full 122-day window were correspondingly lower (evening 56% vs 34%, lenient overall 66% vs 44%; both p < 0.05). After at-risk adjustment, which accounted for retention differences, these group differences disappeared (both p > 0.48). On days when engaged Pain participants used the app, they used it as intensively as engaged Healthy controls.

A further pattern emerged in the weekly trajectories. Among participants who remained active, the Pain group became progressively more engaged than Healthy controls from approximately week 8 onward, with the crossover concentrated in momentary and voluntary check-ins. That is interesting because these assessment types are most dependent on intrinsic motivation. This is consistent with prior evidence that chronic pain patients with greater symptom severity use pain management apps more frequently (Jamison et al. 2017) and with the hypothesis that personal relevance sustains engagement over long timescales, though the present design does not isolate relevance as a causal factor.

Within the Pain group, however, the picture is more nuanced. Indicators of baseline pain burden (pain catastrophizing and pain intensity) emerged as the largest risk factors in the exploratory elastic net, but the model achieved only modest discrimination (AUC 0.57) on a small held-out sample, and baseline predictors collectively accounted for less than a fifth of the variance in temporal coverage. The signal that participants with greater clinical burden may also be those most likely to disengage is therefore best treated as hypothesis-generating and interpreted cautiously. It is, however, not inconsistent with what is currently known. The existing literature points in two directions that are easily mistaken for a contradiction: chronic pain patients with greater symptom severity have been observed to use pain management apps more intensively when active (Jamison et al. 2017), while in longer-duration remote monitoring of mental health populations, more severely affected participants have shown lower retention (Vachon et al. 2019; Zhang et al. 2023). Both patterns can hold simultaneously if severity intensifies within-session engagement but also accelerates eventual disengagement. What is more robust in the present data is the group-level pattern: the Pain group disengaged at roughly twice the rate of Healthy controls, with implications for the representativeness of any dataset built on this kind of design (Inan et al. 2020). Identifying interventions that close this attrition gap, particularly during the first month, when disengagement is concentrated, is a priority for future work. The current data suggest that targeted onboarding for clinical participants, adaptive re-engagement triggered by week-one behavioral signals, and modified incentive structures are all candidates that warrant systematic testing to improve retention in clinical cohorts.

### Voluntary check-ins capture engagement that prompted compliance metrics miss

A quarter of all check-ins were voluntary: self-initiated, uncompensated, and outside any protocol requirement. At the individual level, the median participant’s check-in composition was 26.8% voluntary, and nearly one in six participants maintained voluntary check-ins without a two-week lapse for the full four months. Voluntary rates did not differ between Pain and Healthy participants. Standard compliance metrics, which compute response rates against scheduled prompts, exclude these data by definition.

This matters in two ways. First, it reframes individual-level compliance: a participant classified as 30% compliant under our strict compliance measures (completing both prompted check-in types on fewer than one in three days), may nonetheless have been actively using the app on many additional days through voluntary entries. The total data contribution of such a participant is substantially greater than the prompted compliance rate suggests, and the cumulative dataset substantially larger than scheduled-prompt accounting captures. Second, sustained voluntary self-monitoring in the absence of any external incentive suggests that for a meaningful proportion of participants, the act of tracking was useful in itself rather than merely tolerable. The Soma app’s design using brief momentary check-ins (under 30 seconds) and a personal trends page that visualized participants’ own pain and mood trajectories likely contributed to this, though the present design does not isolate which features mattered.

The broader point is that prompted compliance is one channel of engagement among several, and the field currently lacks a standard way to quantify the others (Torous, Michalak, and O’Brien 2020). Event-contingent and participant-initiated reporting have a long methodological history in diary research (Bolger, Davis, and Rafaeli 2003) but have been underused in EMA, where the dominant design choice has been signal-contingent prompting on a fixed schedule. For digital health protocol design, the implication is that incorporating self-initiated reporting alongside scheduled prompts may yield two distinct benefits: a richer dataset that captures the moments participants themselves consider worth recording, and a measurable proxy for intrinsic engagement that scheduled-prompt compliance cannot provide. This may be particularly valuable in remote monitoring designs intended to run over months or years, where intrinsic motivation likely matters more than protocol-driven prompting for sustained data collection (Inan et al. 2020).

### Design features as candidate supports for sustained engagement

The present study was not designed to isolate which features of the Soma protocol drove the engagement patterns observed, and any account of mechanism must therefore be hypothesis-generating. Several design choices nonetheless warrant explicit mention as candidates for direct testing in future protocols.

Assessment burden is the most empirically defensible candidate. Soma momentary check-ins were completable in under 30 seconds and the evening assessment took approximately three minutes, placing both at the lower end of assessment burden reported in the EMA literature. Experimental evidence indicates that shorter assessments sustain compliance and reduce careless responding more effectively than longer ones, particularly under higher sampling frequencies (Eisele et al. 2022). For protocols extending over months rather than days, in which cumulative burden compounds with every additional second across hundreds of repetitions, item-level brevity may be a more important design lever than is currently reflected in EMA practice.

A second candidate is reciprocity. The Soma trends page gave participants ongoing access to their own pain and mood trajectories, transforming the app from a one-way data collection instrument into one that returned something of personal value in exchange for the data contributed and potentially promoted self-management behaviors. This is consistent with broader evidence that engagement with remote measurement technology is shaped substantially by perceived personal utility (Simblett et al. 2018). The sustained voluntary check-ins reported in the previous subsection are at least consistent with participants finding the act of tracking personally useful, though the present design cannot test this directly. The re-engagement schedule (automatic reminder emails) and modest compensation structure ($0.50 per day, $5 monthly bonus) may also have contributed. Pratap et al. (2020) found that compensation was associated with longer retention in remote digital health studies, but whether any of these features had incremental effects beyond the others cannot be determined here (Pratap et al. 2020).

For digital health protocol design, the broader implication is not that researchers should adopt these specific features but that the design space for long-duration remote monitoring contains more degrees of freedom than is typically tested. Brief assessment items, personal feedback features, graded re-engagement, and sustained modest compensation are all independently manipulable, and systematic testing of their incremental contributions to retention and compliance would meaningfully advance the design of decentralized trials and remote chronic disease monitoring (Inan et al. 2020).

## Limitations

Several limitations should be considered. The analytic sample of 214 participants reflects substantial pre-enrollment attrition (2,943 screened, 429 consented, 256 enrolled, 214 in the final analytic sample), whereby participants who completed each successive step may differ systematically from those who did not. The retained sample was predominantly female (83%) and white (77%), recruited largely through social media and Rhode Island community advertising, limiting generalizability to more diverse populations and settings. The Healthy control group (n = 41) was substantially smaller than the Pain group (n = 173), reflecting Soma’s design as a pain-monitoring platform; this reduced power for group comparisons, and the small acute/subacute subgroup (n = 19) motivated its combination with chronic pain participants for primary analyses.

The app infrastructure did not retain push-notification delivery logs, so all compliance rates are computed against the theoretical maximum number of prompts rather than prompts actually delivered. True prompt-conditional compliance is therefore likely higher than reported, and the at-risk adjustment does not address delivery failures during active periods. Future studies should prioritize server-side delivery logging.

The at-risk adjustment depends on operationalizing disengagement as a 14-day lapse; alternative thresholds would yield somewhat different adjusted rates, though the overall pattern is unlikely to depend on this cutoff. The elastic net classifier was evaluated on a held-out sample of only 39 participants and should be treated as hypothesis-generating.

Voluntary check-ins are interpreted as a channel of intrinsic engagement, but this interpretation is not externally validated; self-initiated entries may also reflect symptom-driven flare-up logging or other behaviors not captured by an intrinsic-engagement framing. The study was purely observational, and feasibility under an active treatment arm may differ. Findings derive from a single platform with one assessment protocol and may not generalize to other long-duration EMA designs. Rolling enrollment between June 2023 and March 2026 means seasonal effects cannot be ruled out, and although the 122-day window is far longer than typical EMA protocols, whether the stable in-protocol compliance plateau would extend further remains untested.

## Conclusion

Long-duration daily EMA-based symptom tracking in a clinical population is feasible, but expectations for what feasibility should look like at this timescale require adjustment compared to more common shorter protocols. Among participants who remained active in this protocol, compliance did not decay across four months. This pattern runs counter to linear extrapolations from short-protocol benchmarks and emerges only when retention and compliance are reported as separate quantities rather than collapsed into a single adherence figure. The bottleneck for long-duration remote symptom monitoring is not that engaged participants tire of the protocol; it is that early disengagement is concentrated in the first month and falls disproportionately on the clinical group. These findings reframe the practical challenge: retention efforts should focus on the first month, when behavioral signals of impending disengagement are already detectable; adherence reporting should separate retention from compliance; and the design space for long-duration remote monitoring, including assessment brevity, personal feedback, and intrinsic engagement channels such as voluntary self-initiated reports, contains more degrees of freedom than current practice has tested. With retention-focused onboarding, separated adherence reporting, and design features that sustain intrinsic engagement, intensive multi-month EMA-based symptom tracking appears feasible at the timescales that digital health, decentralized trials, and longitudinal chronic disease monitoring actually require.

## Data Availability

The analytic code and de-identified summary data supporting the findings of this study will be made publicly available on GitHub upon publication. Prior to publication, data will be made avaiable upon reasonable request to the authors.

## Acknowledgments

The authors wish to thank Madison Corinha, Nova Chen, Phillip Meader Yetter and Claire Dick for their contributions to data collection, recruitment and study support. We also thank the Center for Computation and Visualization (CCV) team at Brown University, especially Brad Roarr, Ellen Duong, and Isabel Restrepo, for their support in developing and maintaining the Soma App.

## Authors’ Contributions

Conceptualization: CZG, FHP; Data curation: PP, CZG; Formal analysis: FHP, CZG; Funding wiacquisition: FHP, CZG; Methodology: FHP, CZG; Resources: FHP, AC; Software: FHP, CZG; Supervision: FHP; Writing – original draft: CZG, FHP; Writing – review & editing: CZG, PP, AC, FHP

## Data & Code Availability

The analytic code and de-identified summary data supporting the findings of this study will be made publicly available on GitHub upon publication.

## Abbreviations

EMA: Ecological Momentary Assessment
KM: Kaplan-Meier
BPI: Brief Pain Inventory
PCS: Pain Catastrophizing Scale
PHQ-8: Patient Health Questionnaire (8-item)
GAD-7: Generalized Anxiety Disorder Scale (7-item)
IQR: Interquartile Range
HR: Hazard Ratio
CI: Confidence Interval

